# Chronic Overlapping Pain Conditions and Nociplastic Pain

**DOI:** 10.1101/2023.06.27.23291959

**Authors:** Keira J.A. Johnston, Rebecca Signer, Laura M. Huckins

## Abstract

Chronic Overlapping Pain Conditions (COPCs) are a subset of chronic pain conditions commonly comorbid with one another and more prevalent in women and assigned female at birth (AFAB) individuals. Pain experience in these conditions may better fit with a new mechanistic pain descriptor, nociplastic pain, and nociplastic type pain may represent a shared underlying factor among COPCs. We applied GenomicSEM common-factor genome wide association study (GWAS) and multivariate transcriptome-wide association (TWAS) analyses to existing GWAS output for six COPCs in order to find genetic variation associated with nociplastic type pain, followed by genetic correlation (linkage-disequilibrium score regression), gene-set and tissue enrichment analyses. We found 24 independent single nucleotide polymorphisms (SNPs), and 127 unique genes significantly associated with nociplastic type pain, and showed nociplastic type pain to be a polygenic trait with significant SNP-heritability. We found significant genetic overlap between multisite chronic pain and nociplastic type pain, and to a smaller extent with rheumatoid arthritis and a neuropathic pain phenotype. Tissue enrichment analyses highlighted cardiac and thyroid tissue, and gene set enrichment analyses emphasized potential shared mechanisms in cognitive, personality, and metabolic traits and nociplastic type pain along with distinct pathology in migraine and headache. We use a well-powered network approach to investigate nociplastic type pain using existing COPC GWAS output, and show nociplastic type pain to be a complex, heritable trait, in addition to contributing to understanding of potential mechanisms in development of nociplastic pain.

## Background

Chronic pain can be defined as pain that persists 3+ months ^1^ and is a main symptom of many conditions as well as being associated with injury and surgery. Chronic pain can also be studied as a complex disease trait as evidenced by large recent genome wide association studies (GWAS) ^2–7^. Recently, the International Association for the Study of Pain (IASP) also redefined pain and ‘chronic primary pain’ codes for the International Classification of Diseases 11^th^ Edition (ICD-11) ^8–10^. More than 1 in 5 US adults experience chronic pain ^11^, and chronic pain is associated with high socioeconomic and quality of life burden ^1,12^, and despite genetic studies into chronic pain as a disease trait, and on conditions where chronic pain is a prominent symptom, mechanisms of development of chronic pain are not fully understood.

Neuropathic and nociceptive pain ^13^ are used to categorize pain and chronic pain according to suspected or confirmed underlying mechanism(s). Neuropathic pain is defined as being caused by lesions or disease in the somatosensory nervous system, and nociceptive pain, designed to directly contrast neuropathic pain, is defined as pain arising from actual or threatened damage to non-neural tissue (in the context of a normally functioning somatosensory nervous system). However, pain experienced in the context of many different chronic pain conditions may not fit with these two descriptors – Chronic Overlapping Pain Conditions (COPCs) ^14–16^ may represent a subset of chronic pain conditions where pain experiences very often diverge from nociceptive/ neuropathic or a mixed nociceptive/ neuropathic pain state. COPCs are a subset of chronic pain conditions commonly comorbid with one another and more prevalent in people assigned female at birth (AFAB). The United States congress and the National Institutes of Health (NIH) listed ten conditions as COPCs; myalgic encephalitis/ chronic fatigue syndrome (ME/CFS), vulvodynia, temporomandibular disorders, irritable bowel syndrome (IBS), interstitial cystitis/ painful bladder syndrome, fibromyalgia, endometriosis, chronic tension-type headache, chronic migraine headache, and chronic low-back pain. In these conditions, there may not be actual or threatened tissue damage or lesion/ disease at the somatosensory nervous system (e.g., fibromyalgia), or if features of nociceptive/ neuropathic pain are present they do not fully capture the pain experience (e.g., chronic low back pain). A proposed third mechanistic pain descriptor, ‘nociplastic pain’, added to IASP terminology ^13^ in 2017 may better capture features of pain in COPCs ^14^.

Nociplastic pain is defined as pain arising from altered nociception in the absence of clear lesion/ disease of the somatosensory nervous system and/or absence of actual or threatened tissue damage. Pain in COPCs, where nociplastic pain could be the underlying mechanism (or main underlying mechanism if a mixed pain state is suspected), may fit better with this description as many COPCs are not associated with neural/ non-neural tissue damage (e.g., fibromyalgia), or if disease and tissue damage are present pain is often non-proportional to tissue damage and can be diffuse throughout the body beyond diseased tissue sites (e.g., endometriosis, chronic low back pain). Nociplastic pain is also associated, compared to nociceptive or neuropathic pain, with greater risk of CNS-related symptoms such as fatigue, changes in cognition and memory, depression, anxiety, and sleep issues as also commonly seen in COPCs ^15^.

These features of COPCs, combined with the fact they are commonly comorbid with one another, may suggest that nociplastic type pain is an underlying shared factor across COPCs. Even in COPCs where a mixed pain state with nociceptive pain (i.e., where tissue damage may be present) such as endometriosis, studies have shown that nociplastic pain contributes significantly to pain experience, and is associated with severity of pain independent of surgical procedures, amount of endometriosis, body mass index, and age ^17^. Previous studies also suggest COPCs could be viewed as a single “lifelong” disease that “manifests in different bodily regions over time” ^18^. There are epidemiological studies of pelvic pain conditions (e.g., MAPP ^19,20^), but there are no GWAS of vulvodynia, chronic pelvic pain, or bladder pain syndrome/ IC. In addition, there are to date no sufficiently large studies with both genotyping and questionnaire data designed specifically to ascertain nociplastic pain (such as the Central Sensitization Inventory ^21^ or Nociplastic-Based Fibromyalgia Features tool ^22^). However, large-scale GWAS for several other COPCs are available, and can be studied as a network to uncover genetic variation associated with nociplastic-type pain as an underlying factor in all COPCs.

Chronic pain conditions can be highly stigmatized ^23–27^ – this is likely even more common in COPCs due to a lack of or disproportionate-to-pain-level presence of tissue or nerve damage ^28–30^, and higher prevalence of COPCs in women and AFAB people ^23,31^. Understanding mechanisms of chronic pain that are not due to, or cannot be fully explained by, nervous system damage or dysfunction or tissue damage (i.e., nociplastic pain) will contribute to legitimizing the pain experience in COPCs. In addition, finding genetic variation associated with nociplastic pain could inform new treatment approaches in COPCs, conditions where many existing pharmaceutical and surgical treatments can be less effective ^32^ or even actively worsen pain ^33^ compared to use in nociceptive/ neuropathic pain.

## Methods

### GWAS summary statistics

GWAS summary statistics for 6 chronic overlapping pain condition (COPC) traits were obtained through publicly available downloads, requests made directly to study authors, or through data request to FinnGenn (release R9, May 2023). Traits, sample sizes, and sources are summarized in Table 1, and include chronic widespread pain ^2^, low back pain ^34^, broad headache ^35^, temporomandibular joint disorder ^36^, and irritable bowel syndrome ^37^. We opted not to include migraine or ME/CFS summary statistics. With ME/CFS ^38^ this is due to sample size being too small for successful completion of the multivariable LDSC step of GenomicSEM common-factor GWAS analysis (Ncase = 427, Ncontrol = 972), and because genotyping in this study was performed using the Illumina Immunochip array (i.e., not whole-genome genotyping, but a specialized assay focused on immune-relevant SNPs). For migraine, a significant proportion of the ‘broad headache’ GWAS case participants are likely migraineurs and/or have both tension-type headache and migraine, therefore including the broad headache GWAS but not migraine GWAS allows for capturing a fuller spectrum of headache and migraine-associated genetic variation, without over-representation of migraine.

**Table 1:** Sources and sample sizes for COPC trait GWAS. See Table S1 for sample and population prevalence estimates and their sources.

| Trait | Acronym | Source | Cases | Controls |
| --- | --- | --- | --- | --- |
| Chronic widespread pain | CWP | Rahman et al 2021<sup>2</sup> | 6914 | 242929 |
| Endometriosis | NA | FinnGenn (R9, May 2023) | 15088 | 107564 |
| Low-back pain | LBP | Suri et al 2021<sup>34</sup> | 49182 | 51629 |
| Broad headache | NA | Meng et al 2018<sup>35</sup> | 74461 | 149312 |
| Temporomandibular joint disorder | TMJ | Jiang et al 2021 (GWAS Catalog)<sup>36</sup> | 217 | 456131 |
| Irritable Bowel Syndrome | IBS | Eijsbouts et al<br>2021 (GWAS<br>Catalog)<sup>37</sup> | 53400 | 433201 |

For each trait excluding endometriosis, GWAS samples are at least partially, and in some cases completely, comprised of UK Biobank participants and samples may overlap –sample overlap is permitted and does not statistically bias GenomicSEM analyses ^39,40^.

### Common factor GWAS

GenomicSEM was used to carry out a common-factor GWAS. First, we prepared (munged) GWAS summary statistics using ‘munge’ function included in GenomicSEM. Sum of effective sample sizes was calculated for low back pain (LBP) and IBS, as these represented GWAS meta-analyses of case-control phenotypes ^41^. Briefly, variants with info values <= 0.9, minor allele frequency (MAF) <= 0.01, that were not SNPs with missing values, that were strand ambiguous, those with duplicated rsIDs, those without a match in the HapMap 3 SNP file used for quality control, and those with mismatched allele labels compared to the HapMap 3 SNP file were removed. Next, multivariable linkage disequilibrium (LD)-score regression was carried out to produce matrices used in the common-factor GWAS step. GenomicSEM ‘sumstats’ function was used for a final preparation step, jointly processing the GWAS summary statistics files for each of the 6 traits included in the analysis. This function merges across all summary statistics using listwise deletion, performs quality control (including backing out logistic betas, checking for allele mismatches, missing data, and duplicate variants), and merges with the reference SNP file. Output from multivariable LD-score regression and from GenomicSEM ‘sumstats’ is taken forward to common factor GWAS using GenomicSEM ‘commonFactorGWAS’ function. Our chosen common-factor model, where the single latent factor represents ‘nociplastic type pain’, is shown in Figure 1.

**Figure 1:**
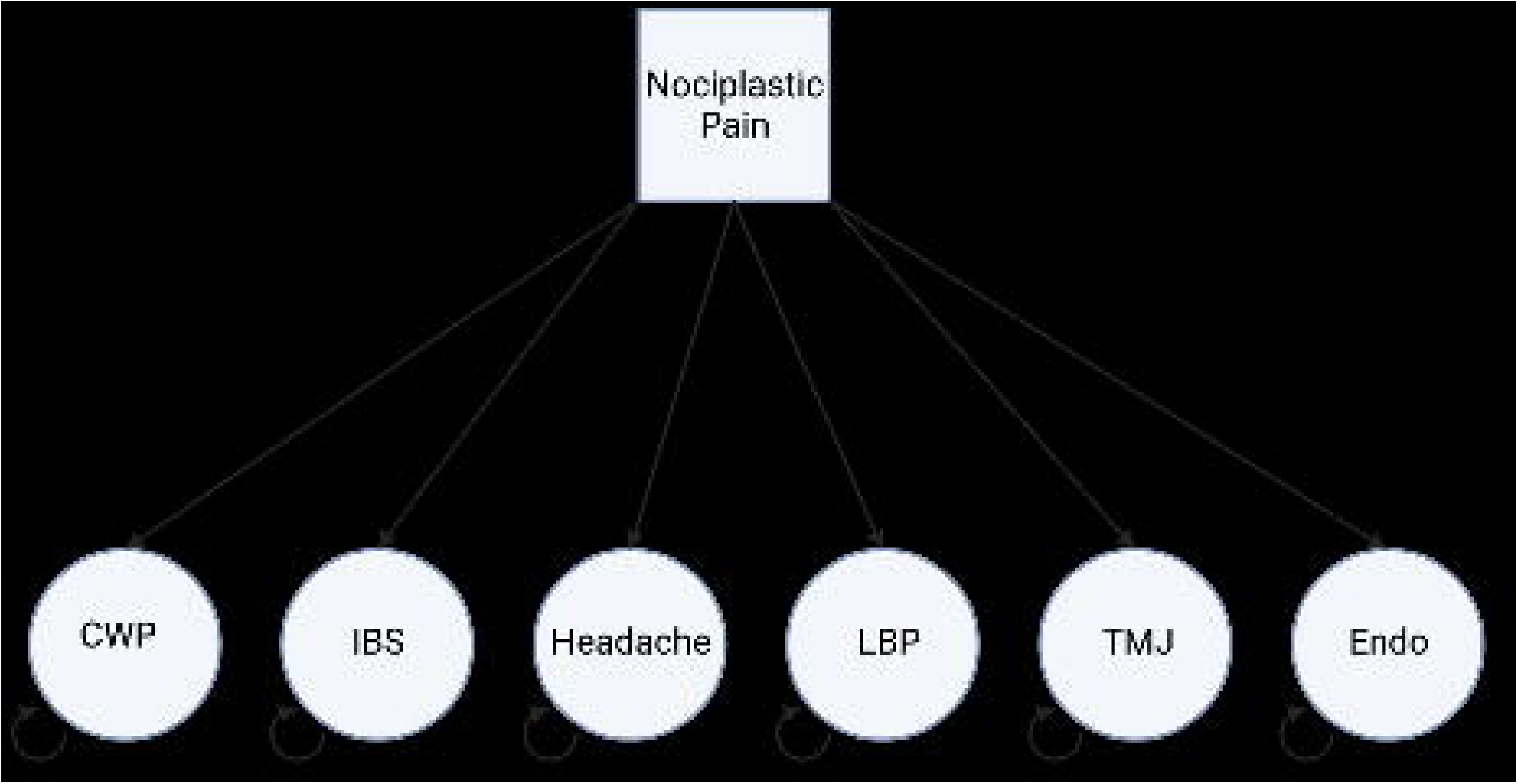
Path diagram of common factor GWAS GenomicSEM model. Values = Standardized Estimate (Standard Error).

First, we fitted the common-factor GWAS model without SNP effects and noted loadings of each trait onto the singular factor (Figure 1). These were significant (Table 2, p < 0.05) and model fitting was successful, so we proceeded to fitting the model including SNP effects with ‘smooth_check=T’. We note that TMJ variance standard error is very large (Figure 1, SE = 3.8), likely due to the small case number in the TMJ GWAS, but factor loading was significant (Table 2, 0.69, p < 0.05) so we elected to include TMJ in this model. Output produced from the above model is equivalent to GWAS summary statistics (magnitude of association between each SNP and a trait of interest) for the latent factor onto which the 6 traits load – which we describe as nociplastic-type pain. Next, we calculate an estimated sample size for the ‘GWAS’ of nociplastic-type pain according to instruction provided on the GenomicSEM github, first reserving SNP results for SNPs with MAF <= 0.4 and => 0.1, and then calculating N̂ according to the formula for each SNP:

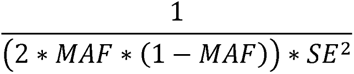

**Equation 1: Formula to estimate per-SNP sample size for SNPs included in common-factor GWAS**

**Table 2:** Standardized common-factor GWAS model factor loadings and p values. SE = standard error.

| Trait | Standardized<br>Estimate | Standardized<br>SE | p-value |
| --- | --- | --- | --- |
| CWP | 0.76 | 0.06 | $1.78 \times 10^{-41}$ |
| IBS | 0.62 | 0.03 | $3.87 \times 10^{-73}$ |
| Broad Headache | 0.73 | 0.04 | $4.56 \times 10^{-81}$ |
| LBP | 0.39 | 0.04 | $5.53 \times 10^{-28}$ |
| TMJ | 0.69 | 0.29 | 0.0188 |
| Endometriosis | 0.57 | 0.06 | $3.34 \times 10^{-20}$ |

Taking the mean of this set of values then gives N̂. For nociplastic-type pain N̂ =578,561.3 (where this sample size is necessary in calculations e.g., LD-score regression analyses for genetic correlation, we use the value 578,561).

### Defining significant, independent SNPs

Common-factor GWAS output (nociplastic type pain summary statistics) was then taken forward and analyzed within FUMA (Functional Mapping and Annotation of Genome-Wide Association Studies) ^42^, a web-based suite of tools for downstream GWAS analyses. Genome-wide significant, independent SNPs were defined using FUMA as SNPs associated with nociplastic pain factor (p < 5×10^-8) and independent from one another (r^2^ < 0.6).

### LD-score Regression

We used ‘ldsc’ ^40,43^ to estimate SNP-heritability of nociplastic-type pain, and to estimate genetic correlation between nociplastic-type pain and three pain-related phenotypes; multisite chronic pain (MCP), a measure of number of chronic pain sites, rheumatoid arthritis, a chronic pain condition that is not usually considered a COPC and where pain for a majority of individuals is likely mainly nociceptive/ inflammatory ^44,45^, and a neuropathic pain phenotype ^46^.

### Multisite chronic pain vs nociplastic-type pain

We obtained summary statistics for GWAS of multisite chronic pain (MCP) ^3^, a general chronic pain phenotype previously found to be heritable, polygenic, and significantly associated with gene expression changes in the brain through download from University of Glasgow Enlighten (research data repository). MCP summary statistics were munged as previously described, using ‘ldsc’ package, as was common factor GWAS output for nociplastic-type pain. We then carried out LD-score regression, estimating genetic correlation between MCP and nociplastic-type pain.

### Rheumatoid arthritis vs nociplastic-type pain

We obtained rheumatoid arthritis GWAS summary statistics from a recent large GWAS (N = 311, 292) ^47^ from GWAS Catalog ^48^. As previously described and again using ‘ldsc’, we calculated genetic correlation between nociplastic type pain and rheumatoid arthritis.

### Neuropathic vs nociplastic-type pain

We obtained GWAS summary statistics for a study on neuropathic pain susceptibility (N_effective_ = 16,311.72) by request to the study authors ^46^, using ‘ldsc’ as previously described to calculate genetic correlation between nociplastic-type pain and neuropathic pain.

### Multivariate Transcriptome-Wide Association Study

An extension of GenomicSEM common-factor GWAS is multivariate (common-factor) TWAS. Here, FUSION TWAS ^49^ output for each of the 6 COPCs serves as input (as GWAS summary statistics did for common-factor GWAS in GenomicSEM common-factor GWAS). We performed TWAS in each of the 6 COPC traits using GWAS summary statistics and the FUSION package and scripts, and using pre-computed predictive models for all 49 Genotype-Tissue Expression project (GTEx) ^50^ v8 tissues ^51^. We used models including genes with significant heritability and with weights calculated using all genetic ancestries, as recommended for typical analyses and to increase sensitivity. FUSION output (TWAS summary statistics for each of the 6 COPCs) was prepared using the GenomicSEM ‘read_fusion’ function, and then taken forward with multivariable LDSC output from previous common-factor GWAS analysis to perform common-factor TWAS.

Significant gene-tissue association findings were defined at the tissue-wide and experiment-wide level, through Bonferroni multiple testing correction across all tests within a tissue and across all genes tested across all tissues respectively.

### Specific and Non-Specific Genes

As part of multivariate GWAS and TWAS analyses, a Q (heterogeneity) value is calculated per SNP-trait association (or per gene-tissue-trait association), indicating degree of heterogeneity (i.e., the proportion of gene expression or SNP association effect that is mediated through pathways other than the shared common factor). Genes specific to nociplastic-type pain were identified through subsetting multivariate TWAS output to include genes in significant (tissue-wide) gene-tissue associations and with non-significant Q p values (i.e., non-significant heterogeneity). Non-specific genes were also identified as above but with significant Q p values. Q p values were Bonferroni-corrected for multiple testing within-tissue. (Qp_Bonferroni_ = Qp/N tests in that tissue).

### Tissue enrichment analyses

We carried out binomial tests of enrichment within our TWAS results to investigate whether certain tissues showed a higher proportion of tissue-wide significant (P_Bonferroni_ < 0.05) associations results than expected by chance, and whether certain tissues showed higher proportion of nominally significant (P < 0.05) associations than expected by chance. Genes tested per tissue are available from FUSION predictor models ^49^.

### Gene-set enrichment analyses

Gene-set enrichment analyses on all tissue wide significant genes, on tissue wide significant and specific genes (Qp_Bonferroni_ > 0.05), and on tissue wide significant non-specific genes (Qp_Bonferroni_ < 0.05) was carried out using FUMA, with all genes tested in multivariate TWAS and with recognized Ensembl gene ID as background (N = 26455 genes).

## Results

### Common-factor GWAS

We fitted a common factor GWAS model (including individual SNP effects), using data for six COPC traits (CWP, LBP, endometriosis, TMJ, IBS, and broad headache). This model estimates the size of association between SNPs and a latent common factor (nociplastic type pain), producing results effectively equivalent to a standard GWAS of this unmeasured latent factor. We found a total of 663 SNPs across fifteen GWAS genomic risk loci significantly associated with nociplastic-type pain (p < 5 x 10^-8, Figure 2), consisting of 24 independent SNPs. The majority of these SNPs (18/24) have not been previously associated with pain-related traits (Table S2).

**Figure 2:**
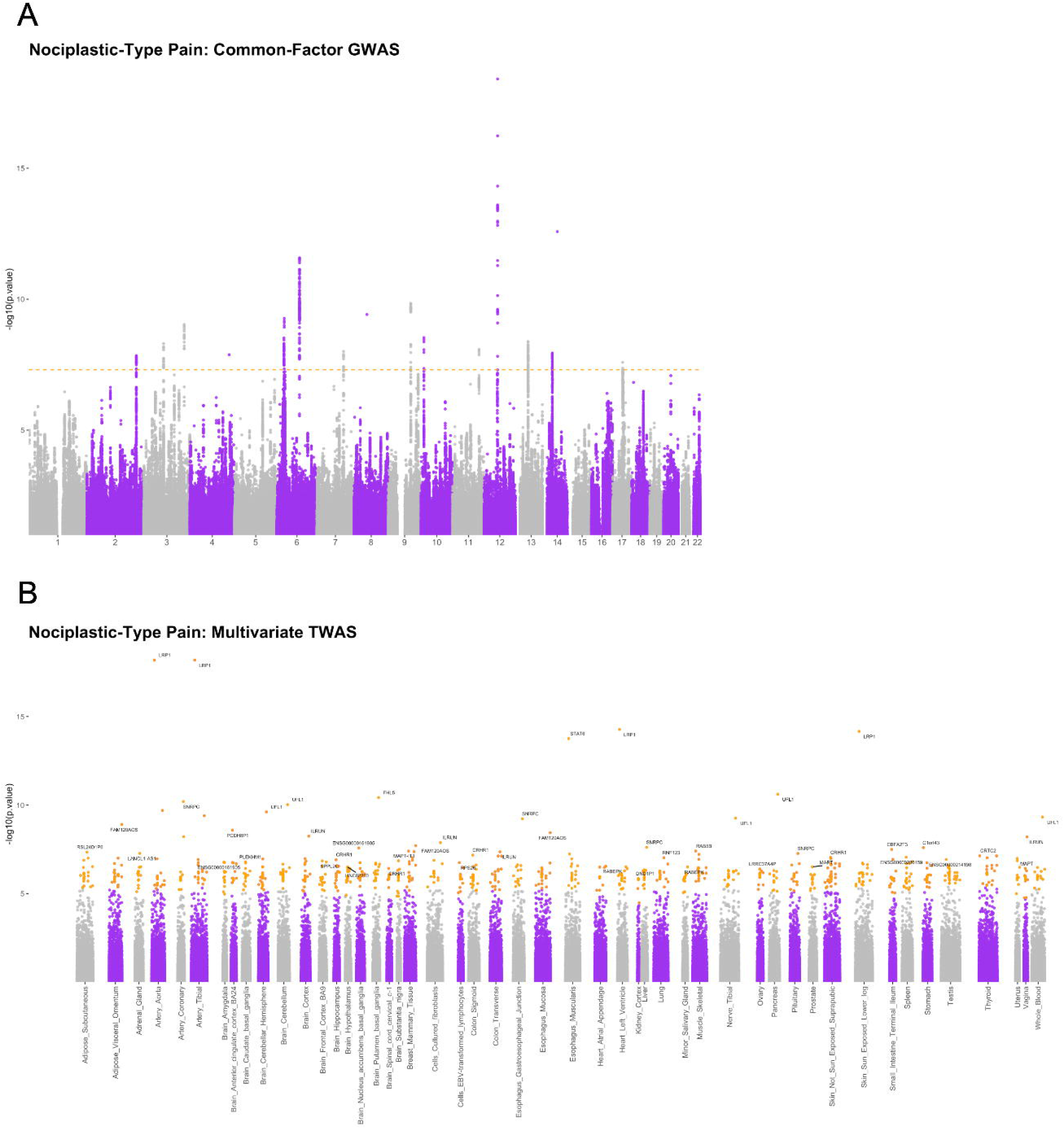
Manhattan plot of nociplastic-type pain common-factor GWAS and multivariate TWAS outputs. A: Nociplastic-type pain – common-factor GWAS Manhattan plot. Dotted line = genome-wide p value significance threshold (-log_10_(5 x 10^-8^)). B: Nociplastic-type pain – multivariate TWAS Manhattan plot. X-axis labels = GTEx v8 tissue, points shown in orange = gene-tissue associations significant after Bonferroni adjustment within that tissue, labeled points = genes with most significant association within that tissue.

### Heritability and Genetic Correlation

We estimated the SNP-based heritability of the nociplastic type pain trait using output from our GenomicSEM common-factor GWAS. We also estimated genetic correlation between nociplastic type pain and another complex chronic pain trait, multisite chronic pain, between nociplastic type pain and RA, a disease where chronic pain is a common symptom but is likely associated with nociceptive rather than nociplastic pain mechanisms ^44,45^, and between nociplastic-type pain and neuropathic pain. Nociplastic-type pain was found to be significantly heritable (liability scale SNP-h2 = 0.025, SE = 0.0014), and was significantly genetically correlated with MCP (rg = 0.92, SE= 0.04) and to a much lesser degree with rheumatoid arthritis (rg = 0.18, SE = 0.04). Neuropathic and nociplastic-type pain were also significantly genetically correlated (rg = 0.79, SE = 0.099).

The LD-score regression intercept values when calculating these genetic correlations was close to 1 (0.97-1.03), suggesting the majority of genomic inflation captured by lambda GC (estimates ranging 1.0025-1.3) is due to polygenicity rather than population structure.

### Multivariate TWAS

To explore nociplastic type pain at the transcriptomic level and uncover tissue-level gene expression relevant to this trait, we carried out a common-factor TWAS analysis of 6 COPC traits using GenomicSEM. We found 819 tissue-wide significant gene-tissue associations consisting of 127 unique genes, across all 49 tested GTEx tissues. As part of multivariate GWAS and TWAS analyses with GenomicSEM, a Q (heterogeneity) value is calculated per association. This value indicates degree of heterogeneity (i.e., the proportion of gene expression or SNP association effect that is mediated through pathways other than the shared common factor, across the traits included in the model that load onto the common factor). Specific genes were identified through subsetting multivariate TWAS output to include genes in significant (tissue-wide) gene-tissue associations and with non-significant Q p values (i.e., non-significant heterogeneity). Non-specific genes were also identified as above but with significant Q p values. Q p values were Bonferroni-corrected for multiple testing within-tissue. (Qp_Bonferroni_ = Qp/N tests in that tissue).

450 of the 819 tissue-wide significant gene-tissue associations showed significant heterogeneity (Qp_Bonferroni_ < 0.05), and the remaining 369 did not (Qp_Bonferroni_ > 0.05). Twenty-eight unique genes showed significant heterogeneity across all tested tissues (Qp_Bonferroni_< 0.05), and 94 showed non-significant heterogeneity (Qp_Bonferroni_ > 0.05). Five genes showed both significant and non-significant heterogeneity depending on tissue, and were excluded from gene-set enrichment tests of specific/ heterogeneous genes.

### Tissue enrichment

To determine whether specific tissues showed more or fewer significant TWAS results than expected by chance, we carried out a series of binomial tests for enrichment of significant TWAS results within each tested tissue. We found that 9 of the 49 tested tissues showed a different proportion of significant gene-tissue TWAS findings than expected by chance. Cultured fibroblasts, atrial appendage of the heart, tibial nerve, and thyroid were enriched for significant associations, whereas amygdala, substantia nigra, Epstein-Barr-virus-transformed lymphocytes, terminal ileum of the small intestine and vagina showed significantly fewer significant associations than expected. At the nominal significance level (unadjusted P < 0.05), two tissues showed a different proportion of associations than expected by chance – whole blood and skeletal muscle were enriched for significant (unadjusted P < 0.05) associations.

### Gene-set enrichment

To explore potential mechanisms in nociplastic pain that are shared with other complex traits of interest, we performed three sets of gene-set enrichment tests using FUMA. First, we used all unique tissue-wide significant gene findings from multivariate TWAS, then a subset of those findings that showed significant heterogeneity (non-specific genes) and finally a subset with non-significant heterogeneity (specific genes).

### Tissue-wide significant genes

Of 127 unique genes, 120 had a recognized ensemble gene ID within FUMA and were included in analyses. Eight positional gene sets were enriched (adjusted p < 0.05) for nociplastic-type-pain associated genes; chr17q21, chr3p21, chr9q33, chr16q22, chr12q13, chr2q34, chr4q33, and chr1q21. Kyoto Encyclopedia of Genes and Genomes (KEGG) pathway gene set nitrogen metabolism was also enriched for nociplastic pain genes, as was chemical and genetic perturbation gene set ‘SU_LIVER’ (genes specifically upregulated in the liver). A total of 44 GWAS Catalog traits including cognitive function (adjusted p = 1.76×10^-10^), extremely high intelligence (adjusted p = 3.3×10^-9^), sleep duration (adjusted p = 5.95×10^-9^), and headache (adjusted p = 1.10×10^-8^) were enriched for nociplastic-type pain genes (Table S3, Figure 3A). Additional gene sets enriched for nociplastic pain genes including microRNA target sets, transcription factor target sets, computational gene sets and cancer gene modules can be found in Table S4.

**Figure 3:**
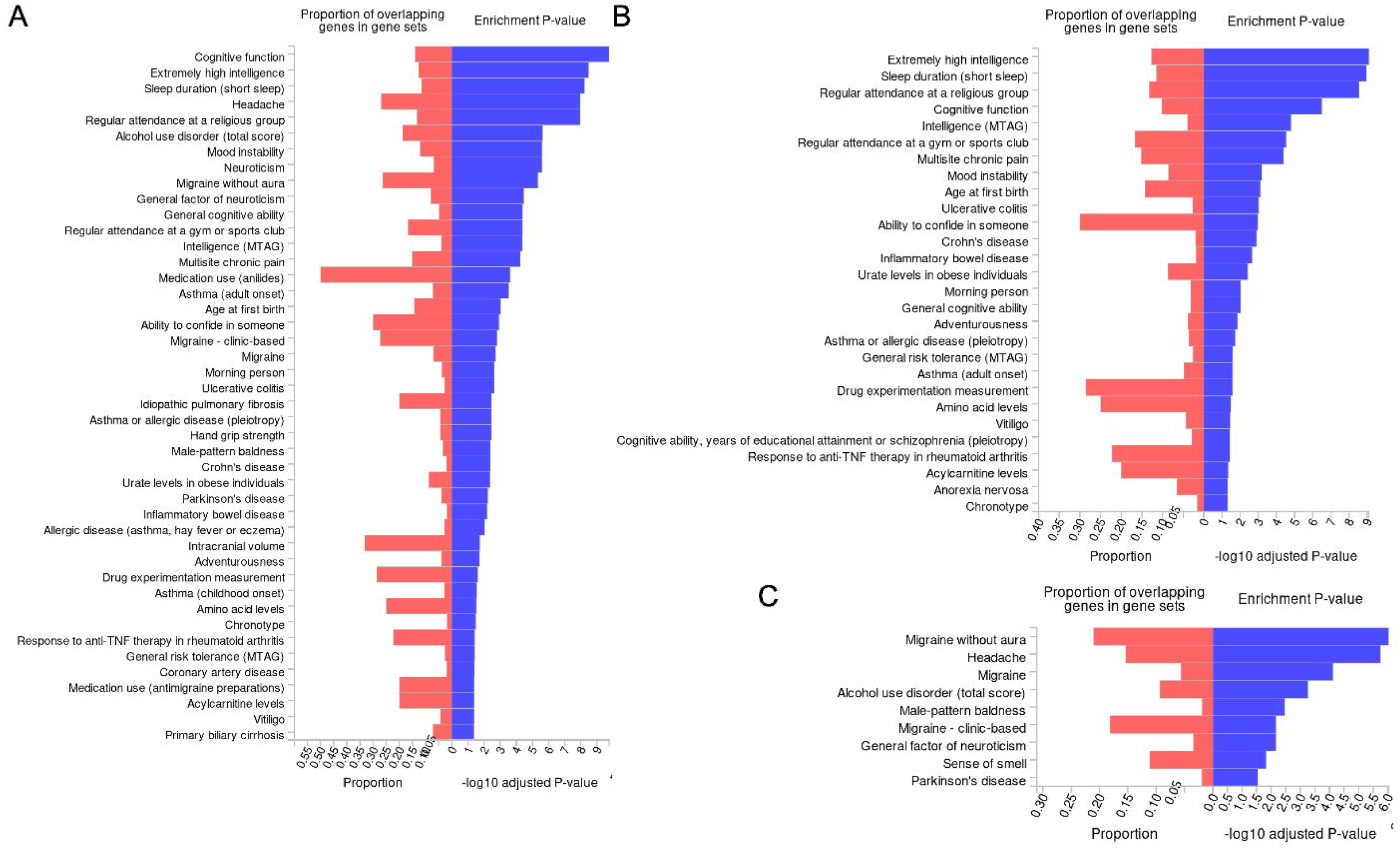
GWAS catalog trait gene set enrichments. A: Gene sets enriched for all nociplastic-type pain genes found in multivariate TWAS analysis, B: Gene sets enriched for specific nociplastic type pain genes, Panel C: Gene sets enriched for non-specific nociplastic type pain genes.

### Specific genes

90 of 94 tissue-wide significant genes without significant heterogeneity across all tissues where a significant association was found, and with an ensembl gene ID recognized within FUMA were included in this analysis. We found seven positional gene sets to be enriched for specific nociplastic pain genes, including chr3p21, chr9q33, chr16q22, chr2q34, chr4q33, chr12q13, and chr1q21. Chemical and genetic perturbation gene set ‘SU_LIVER’ (adjusted p = 4.45×10^-3^) was enriched for specific nociplastic pain genes, as was nitrogen metabolism (adjusted p = 0.01), and the hallmark gene set fatty acid metabolism (adjusted p = 0.018). Additional gene sets enriched for specific nociplastic pain genes are listed at Table S5. We found 28 GWAS Catalog traits enriched for specific nociplastic pain genes (Table S6, Figure 3B), including extremely high intelligence (adjusted p = 8.02×10^-10^), sleep duration (adjusted p = 1.09×10^-9^), regular attendance at a religious group (adjusted p = 2.77×10^-9^), and cognitive function (adjusted p = 3.08×10^-7^).

### Non-specific genes

25 of 28 tissue-wide significant genes with significant heterogeneity across tissues where a significant association was found, and with an ensembl gene ID recognized within FUMA were included in this analysis. Positional gene sets chr17q21, and chr6q16 were enriched for these non-specific nociplastic pain genes, as were several cancer gene neighborhoods (Table S7) and 9 GWAS Catalog traits including migraine without aura (adjusted p = 9.31×10^-7^), headache (adjusted p = 1.79×10^-6^), and migraine (adjusted p = 7.63×10^-5^) (Table S8, Figure 3C).

There was no overlap in gene sets enriched for specific and non-specific genes, and migraine and headache gene sets were enriched for non-specific nociplastic-type pain genes only.

## Discussion

We carried out common-factor GWAS and TWAS analyses incorporating six COPCs to investigate genetic variation associated with nociplastic type pain, a mechanistic pain descriptor and type of pain likely important across COPCs. This method allows us to find genetic variation associated with nociplastic type pain in the absence of a dataset where genotype data and phenotype information on nociplastic type pain are available together. Furthermore, if nociplastic type pain is a shared factor among COPCs, then our findings are likely relevant to COPCs where GWAS are currently unavailable and/ or could not be included here, such as interstitial cystitis, vulvodynia, and ME/CFS. We found that nociplastic type pain is a moderately heritable trait (observed SNP-h2 0.025) with similar SNP heritability to rheumatoid arthritis (RA liability scale SNP-h2 = 0.07, observed SNP-h2 = 0.035). Our findings indicate that nociplastic type pain is a complex, polygenic trait, and we found 24 independent SNPs significantly associated with this trait.

### Nociplastic type pain is genetically correlated with rheumatoid arthritis and MCP

We observed extremely high genetic correlation between nociplastic type pain and MCP (rg = 0.92). Individuals with a non-zero trait value for MCP could as a group be majority composed of individuals with COPCs – to assess this we carried out a series of Fisher’s exact tests on counts of COPC ICD10 code occurrences within and outside of MCP ‘cases’ (MCP trait value >=1) in UK Biobank (see Table S9). We found that for ICD10 category codes for COPCs that were available in UK Biobank (back pain, post-viral fatigue (ME/CFS), migraine, IBS, endometriosis and tension headache), all were significantly overrepresented in MCP cases compared to those without chronic pain (MCP trait value 0), with the most over-represented COPC being fibromyalgia (OR 4.48), considered a prototypical nociplastic pain condition. However, we also tested for enrichment of rheumatoid arthritis (a non-COPC pain condition) cases, and again found significant enrichment (Table S9). This suggests high phenotypic overlap in COPCs and MCP in UK Biobank cannot fully explain high genetic correlation values of our nociplastic pain factor and MCP.

Another explanation for high nociplastic pain factor-MCP genetic correlation may be that a large amount of genetic variation associated with the main characteristic of MCP (increasing number of sites of chronic pain) is also captured in a GWAS/ GWAS-equivalent analysis of nociplastic type pain – a recent paper outlining clinical criteria in assessing and grading nociplastic pain lists regional (as opposed to discrete) location and spread of pain as a key characteristic of nociplastic pain ^52^. Additionally, diagnostic criteria in RA include (chronic) pain in at least 2 or more large joints, which may generate the significant phenotypic overlap between RA and MCP in UK Biobank – however in RA this ‘increasing number of sites’ characteristic is driven by disease-specific processes, resulting in low genetic correlation between RA and nociplastic pain factor, and between RA and MCP.

Neuropathic pain was also relatively highly genetically correlated with nociplastic-type pain (rg = 0.79) – however this rg value is significantly less than 1, indicating a significant portion of genetic variation is distinct between these two pain traits. In addition, a high degree of genetic correlation between nociplastic type and neuropathic pain, to a greater extent than rg with nociceptive pain types, could be expected. In their recent review on nociplastic pain, Fitzcharles et al emphasize not only how common mixed pain states are (pain with nociceptive/neuropathic/nociplastic features), but that neuropathic features in hip and knee arthritis and low back pain observed in studies predating the concept of nociplastic pain likely represent nociplastic etiology ^14^.

Neuropathic features of pain in various rheumatic diseases have also been suggested to actually represent nociplastic pain ^53^.

### Traits enriched for specific and non-specific nociplastic-type pain genes indicate distinct pathology of migraine and headache

Gene set enrichment results showed certain genes significantly contribute to variation in nociplastic pain phenotype, but that a large amount of their influence on COPC traits is not mediated by nociplastic type pain (non-specific genes i.e., genes with significant heterogeneity in genomicSEM analyses). GWAS trait gene sets enriched for such genes included migraine and headache, suggesting involvement of factors outside of nociplastic pain in these traits. Non-nociplastic-type-pain-specific genes were also enriched in trait gene sets for male-pattern baldness (androgenic alopecia) – alopecia, including androgenic alopecia, has been previously associated with drugs that block CGRP (calcitonin gene related peptide), a treatment for acute migraine and migraine prevention ^54^.

### Potential metabolic and liver related pathology in nociplastic pain

GWAS trait gene sets enriched for specific nociplastic type pain genes included amino acid and acylcarnitine levels. Changes in amino acid levels are associated with fibromyalgia, migraine, osteoarthritis and complex regional pain syndrome ^55^. High acylcarnitine levels can indicate disorders in fatty acid metabolism, and diets high in certain fatty acids have been associated with increased allodynia in rodent models, and associated with human chronic pain conditions ^56^. Lipids generally are also involved in acute and chronic inflammation ^57^ and changes to acylcarnitine metabolism are observed in dementia, certain cancers, heart failure and coronary artery disease ^58^.

Changes to circulating lipids have also been observed in COPCs including fibromyalgia, headache, migraine, TMD, low back pain and IBS ^59^, and cholesterol metabolism in microglia has been linked to neuropathic pain in a rodent model ^60^. In addition, certain drug therapies used in treatment for systemic lupus erythematosus and rheumatoid arthritis can disrupt lipid metabolism ^61^, and experiencing chronic pain is associated with changes in diet that can result in changes in lipid profiles ^62^.

Genes specifically upregulated in liver tissue were also enriched for specific nociplastic type pain genes. Gene findings from TWAS in this study in theory most likely represent gene expression mediating the relationship between genotype and trait – in other words gene expression that is likely genetically regulated and that occurs before development of nociplastic pain. Therefore, changes in the liver associated with nociplastic pain may develop prior to nociplastic pain development.

Curcuminoids (components of turmeric) have been previously investigated in treatment of neuropathic pain, and previous studies found that a possible mechanism of action in alleviating neuropathic pain was via modulating nitrogen metabolism ^63^. Although ME/CFS summary statistics were not used in this analysis, changes in nitrogen metabolism have been implicated in this condition ^64^, and our findings suggest these metabolic changes may not be unique to ME/CFS but instead shared across COPCs through influence on nociplastic pain in particular. Those with ME/CFS have also been found to have altered lipid, acylcarnitine, and amino acid levels compared to non-ME/CFS controls ^65^.

### Positional gene sets enriched for nociplastic type pain genes suggest immune and musculoskeletal factors

Positional gene sets associated with specific nociplastic pain genes have also been previously linked to COVID-19 susceptibility and severity (chr3p21, ^66^), schizophrenia and bipolar disorder (chr3p21, ^67^), and Alzheimer disease in an African American cohort (chr3p21, ^7^). Regions in chr9q33 are frequently deleted in certain cancers ^68,69^, and chr16q22 has been previously associated with a rare duplication syndrome accompanied by varying psychiatric disorder symptoms ^70^, and with schizophrenia ^71^. A type of syndactyly (involving fusion of digits and toes) has been associated with chr2q34 ^72^, along with autoimmunity, amyotrophic lateral sclerosis and schizophrenia ^73^, and age-related degeneration in the lumbar spine ^74^. Variants in the chr12q13 region have been associated with childhood obesity ^75^ and asthma ^76,77^. Finally, chr1q21 has been previously implicated in GWAS of circulating interleukin 6 levels ^78^. These findings suggest shared immune and musculoskeletal related etiology in these phenotypes and nociplastic type pain. Other recent studies in the context of pain and long COVID have found that infection can potentially trigger and/or exacerbate existing painful conditions, particularly COPCs including fibromyalgia and ME/CFS, and that those with an existing COPC were more likely to develop long COVID, again suggesting possible immune differences in COPCs ^79^.

In contrast, positional gene sets enriched for non-specific nociplastic pain genes included chr17q21, where duplication and deletion have previously been associated with syndromes involving distinctive craniofacial features, developmental delay, and cardiac symptoms (Online Mendelian Inheritance in Man (OMIM) accession: 610443, 613533), and genes at chr6q16 with cardiac phenotypes ^80^ and cluster headache and migraine ^81^.

### Tissue enrichment of nociplastic type pain genes highlights thyroid, heart, and tibial nerve involvement

Hypothyroidism can both cause pain and worsen pain experienced as part of comorbid chronic pain conditions. Autoimmune disorders including autoimmune hypothyroidism may also be misdiagnosed as fibromyalgia ^82–84^. Enrichment of nociplastic type pain gene associations in thyroid may therefore indicate high levels of thyroid involvement in COPCs, or extensive presence of individuals with comorbid (or misdiagnosed) hypothyroidism among COPC GWAS participants, or both.

Atrial appendages of the heart (left and right) are small pouches located on the front upper surface of the right atrium, and anterior to the left atrium and parallel to the left pulmonary veins. Atrial fibrillation, the most common abnormal heart rhythm in adults, can lead to formation of clots (most commonly in the left atrial appendage) and subsequent stroke ^85–87^. Electrocardiogram abnormalities, particularly atrial fibrillation, have been linked to chronic pain ^88,89^, and with physiological stress associated with chronic illness and major surgery ^90,91^. Chronic pain has also been associated with higher risk of myocardial infarction, death due to cardiovascular event, heart failure, and stroke ^92^. Our findings may suggest this atrial fibrillation in particular could be common in nociplastic pain and COPCs, increasing risk for stroke in these patient populations.

The tibial nerve is one of two terminal branches of the sciatic nerve, providing motor and sensory innervation to almost all of the posterior foot and leg. An ultrasound study comparing participants with fibromyalgia and controls found significant increased cross-sectional area in several nerves, including tibial nerve ^93^. Tarsal tunnel syndrome, a nerve entrapment syndrome analogous to carpal tunnel syndrome in the wrist, was also found to be more common in fibromyalgia patients ^94^ – while causes of tarsal tunnel syndrome are likely multifactorial, enlarged tibial nerve diameter may contribute to this nerve entrapment. Neuromodulation involving the tibial nerve (e.g., through percutaneous tibial nerve stimulation) has also been investigated in the treatment of a range of pelvic pain disorders, including IBS, dysmenorrhea, and bladder pain syndrome ^95^. One caveat is that tibial nerve is the only peripheral nerve tissue sampled in GTEx – other peripheral nerve tissues, potentially also representing therapeutic targets in neuromodulation for chronic pain, could potentially be enriched for nociplastic pain gene expression.

## Conclusions

COPCs are a subset of chronic pain conditions that are commonly comorbid. A third mechanistic pain descriptor, nociplastic pain, may best represent the pain experience of those with COPCs, where tissue and/or nerve damage is often not present. Using existing COPC GWAS data and a network-informed genomics approach, GenomicSEM, we found genetic variation at the SNP, gene expression, and gene-set level associated with nociplastic type pain. We also explored genetic overlap with other chronic pain phenotypes, tissue enrichment of nociplastic pain genes, and differences in genes specific and non-specific to nociplastic-type pain. Our findings indicate distinct pathology in migraine and headache compared to other COPCs and link this distinct pathology with traits such as Parkinson disease, sense of smell, and androgenic alopecia, as well as provide unique genes associated with this pathology. We demonstrate various degrees of genetic overlap between nociplastic type pain and three different pain traits, showing highest overlap with a trait capturing increasing number of pain sites on the body (a key characteristic of nociplastic pain in the literature), and lowest with rheumatoid arthritis, a chronic pain condition where pain mechanisms are considered mostly nociceptive. We also find tissue enrichment relevant to chronic pain comorbidities including stroke and therapies such as peripheral nerve stimulation. Our findings contribute to further understanding mechanisms of nociplastic type pain, and indicate this type of pain is important across COPCs.

## Data Availability

All data produced in the present study are available upon reasonable request to the authors

## Supplemental Information

Supplemental information includes 10 tables.

## Acknowledgements

KJAJ is supported by NIMH (R01MH118278; R01MH124839). RS and LMH are supported by NIMH (R01MH118278; R01MH124839) and NIEHS (R01ES033630). Data associated with UK Biobank approved application number 18177 was used in this study. We thank the study authors who provided their GWAS summary statistics directly to us.

## Web Resources

FUMA: https://fuma.ctglab.nl/

FUSION and genes tested per tissue: http://gusevlab.org/projects/fusion/#gtex-v8-multi-tissue-expression

GenomicSEM: https://github.com/GenomicSEM/GenomicSEM

GWASCatalog: https://www.ebi.ac.uk/gwas/

LDSC: https://github.com/bulik/ldsc

## Data and Code Availability

Individual-level data for UK Biobank is available upon approved application to UK Biobank. Code and resources to implement FUSION, GenomicSEM and LDSC are available at the provided web addresses.

Common-factor GWAS and multivariate TWAS summary statistics produced as part of this study are available for download https://doi.org/10.5281/zenodo.8117582

GWAS summary statistics for broad headache, LBP, and neuropathic pain may be available upon request to the respective study authors.

GWAS summary statistics for endometriosis are available from https://www.finngen.fi/en/access_results

GWAS summary statistics for CWP are available from https://zenodo.org/record/4459546

GWAS summary statistics for IBS are available from http://ftp.ebi.ac.uk/pub/databases/gwas/summary_statistics/GCST90044001-GCST90045000/GCST90044107/

GWAS summary statistics for TMJ are available from http://ftp.ebi.ac.uk/pub/databases/gwas/summary_statistics/GCST90016001-GCST90017000/GCST90016564/

## References

1. Mills, S.E.E., Nicolson, K.P., and Smith, B.H. (2019). Chronic pain: a review of its epidemiology and associated factors in population-based studies. Br. J. Anaesth. 123, e273– e283. 10.1016/j.bja.2019.03.023.

2. Rahman, M.S., Winsvold, B.S., Chavez Chavez, S.O., Børte, S., Tsepilov, Y.A., Sharapov, S.Z., HUNT All-In Pain, Aulchenko, Y.S., Hagen, K., Fors, E.A., et al. (2021). Genome-wide association study identifies *RNF123* locus as associated with chronic widespread musculoskeletal pain. Ann. Rheum. Dis. 80, 1227–1235. 10.1136/annrheumdis-2020-219624.

3. Johnston, K.J.A., Adams, M.J., Nicholl, B.I., Ward, J., Strawbridge, R.J., Ferguson, A., McIntosh, A.M., Bailey, M.E.S., and Smith, D.J. (2019). Genome-wide association study of multisite chronic pain in UK Biobank. PLOS Genet. 15, e1008164. 10.1371/journal.pgen.1008164.

4. Tsepilov, Y.A., Freidin, M.B., Shadrina, A.S., Sharapov, S.Z., Elgaeva, E.E., Zundert, J. van, Karssen, L.С., Suri, P., Williams, F.M.K., and Aulchenko, Y.S. (2020). Analysis of genetically independent phenotypes identifies shared genetic factors associated with chronic musculoskeletal pain conditions. Commun. Biol. 3, 329. 10.1038/s42003-020-1051-9.

5. Johnston, K.J.A., Ward, J., Ray, P.R., Adams, M.J., McIntosh, A.M., Smith, B.H., Strawbridge, R.J., Price, T.J., Smith, D.J., Nicholl, B.I., et al. (2021). Sex-stratified genome-wide association study of multisite chronic pain in UK Biobank. PLOS Genet. 17, e1009428. 10.1371/journal.pgen.1009428.

6. Farrell, S.F., Campos, A.I., Kho, P.-F., de Zoete, R.M.J., Sterling, M., Rentería, M.E., Ngo, T.T., and Cuéllar-Partida, G. (2021). Genetic basis to structural grey matter associations with chronic pain. Brain J. Neurol. 144, 3611–3622. 10.1093/brain/awab334.

7. Ghani, M., Reitz, C., Cheng, R., Vardarajan, B.N., Jun, G., Sato, C., Naj, A., Rajbhandary, R., Wang, L.-S., Valladares, O., et al. (2015). Association of Long Runs of Homozygosity With Alzheimer Disease Among African American Individuals. JAMA Neurol. 72, 1313– 1323. 10.1001/jamaneurol.2015.1700.

8. Nicholas, M., Vlaeyen, J.W.S., Rief, W., Barke, A., Aziz, Q., Benoliel, R., Cohen, M., Evers, S., Giamberardino, M.A., Goebel, A., et al. (2019). The IASP classification of chronic pain for ICD-11: chronic primary pain. Pain 160, 28–37. 10.1097/j.pain.0000000000001390.

9. Treede, R.-D., Rief, W., Barke, A., Aziz, Q., Bennett, M.I., Benoliel, R., Cohen, M., Evers, S., Finnerup, N.B., First, M.B., et al. (2019). Chronic pain as a symptom or a disease: the IASP Classification of Chronic Pain for the International Classification of Diseases (ICD-11). Pain 160, 19–27. 10.1097/j.pain.0000000000001384.

10. IASP Announces Revised Definition of Pain Int. Assoc. Study Pain IASP. https://www.iasp-pain.org/publications/iasp-news/iasp-announces-revised-definition-of-pain/.

11. Yong, R.J., Mullins, P.M., and Bhattacharyya, N. (2022). Prevalence of chronic pain among adults in the United States. PAIN 163, e328. 10.1097/j.pain.0000000000002291.

12. Global, regional, and national burden of 12 mental disorders in 204 countries and territories, 1990–2019: a systematic analysis for the Global Burden of Disease Study 2019 (2022). Lancet Psychiatry 9, 137–150. 10.1016/S2215-0366(21)00395-3.

13. Terminology | International Association for the Study of Pain Int. Assoc. Study Pain IASP. https://www.iasp-pain.org/resources/terminology/.

14. Fitzcharles, M.-A., Cohen, S.P., Clauw, D.J., Littlejohn, G., Usui, C., and Häuser, W. (2021). Nociplastic pain: towards an understanding of prevalent pain conditions. The Lancet 397, 2098–2110. 10.1016/S0140-6736(21)00392-5.

15. Maixner, W., Fillingim, R.B., Williams, D.A., Smith, S.B., and Slade, G.D. (2016). Overlapping Chronic Pain Conditions: Implications for Diagnosis and Classification. J. Pain 17, T93–T107. 10.1016/j.jpain.2016.06.002.

16. Schrepf, A., Phan, V., Clemens, J.Q., Maixner, W., Hanauer, D., and Williams, D.A. (2020). ICD-10 Codes for the Study of Chronic Overlapping Pain Conditions in Administrative Databases. J. Pain 21, 59–70. 10.1016/j.jpain.2019.05.007.

17. Till, S.R., Schrepf, A., Clauw, D.J., Harte, S.E., Williams, D.A., and As-Sanie, S. (2023). Association Between Nociplastic Pain and Pain Severity and Impact in Women With Chronic Pelvic Pain. J. Pain, S1526–5900(23)00369-3. 10.1016/j.jpain.2023.03.004.

18. Harte, S.E., Harris, R.E., and Clauw, D.J. (2018). The neurobiology of central sensitization. J. Appl. Biobehav. Res. 23, e12137. 10.1111/jabr.12137.

19. Schrepf, A., Gallop, R., Naliboff, B., Harte, S.E., Afari, N., Lai, H.H., Pontari, M., McKernan, L.C., Strachan, E., Kreder, K.J., et al. (2022). Clinical Phenotyping for Pain Mechanisms in Urologic Chronic Pelvic Pain Syndromes: A MAPP Research Network Study. J. Pain 23, 1594–1603. 10.1016/j.jpain.2022.03.240.

20. Jr, L., Da, W., Ms, L., Dj, C., Bd, N., Na, R. A, van B. S S., Aj, S., Lv, R., et al. (2014). The MAPP research network: design, patient characterization and operations. BMC Urol. 14. 10.1186/1471-2490-14-58.

21. Neblett, R., Cohen, H., Choi, Y., Hartzell, M., Williams, M., Mayer, T.G., and Gatchel, R.J. (2013). THE CENTRAL SENSITIZATION INVENTORY (CSI): ESTABLISHING CLINICALLY-SIGNIFICANT VALUES FOR IDENTIFYING CENTRAL SENSITIVITY SYNDROMES IN AN OUTPATIENT CHRONIC PAIN SAMPLE. J. Pain Off. J. Am. Pain Soc. 14, 438–445. 10.1016/j.jpain.2012.11.012.

22. GhavidelLJParsa, B., Bidari, A., Atrkarroushan, Z., and Khosousi, M. (2021). Implication of the Nociplastic Features for Clinical Diagnosis of Fibromyalgia: Development of the Preliminary NociplasticLJBased Fibromyalgia Features (NFF) Tool. ACR Open Rheumatol. 4, 260–268. 10.1002/acr2.11390.

23. Perugino, F., De Angelis, V., Pompili, M., and Martelletti, P. (2022). Stigma and Chronic Pain. Pain Ther. 10.1007/s40122-022-00418-5.

24. Bean, D.J., Dryland, A., Rashid, U., and Tuck, N.L. (2022). The Determinants and Effects of Chronic Pain Stigma: A Mixed Methods Study and the Development of a Model. J. Pain 23, 1749–1764. 10.1016/j.jpain.2022.05.006.

25. Collier, R. (2018). “Complainers, malingerers and drug-seekers” — the stigma of living with chronic pain. Can. Med. Assoc. J. 190, E204–E205. 10.1503/cmaj.109-5553.

26. De Ruddere, L., and Craig, K.D. (2016). Understanding stigma and chronic pain: a-state-of-the-art review. Pain 157, 1607–1610. 10.1097/j.pain.0000000000000512.

27. Quintner, J. (2020). Why Are Women with Fibromyalgia so Stigmatized? Pain Med. 21, 882–888. 10.1093/pm/pnz350.

28. Wakefield, E.O., Belamkar, V., Litt, M.D., Puhl, R.M., and Zempsky, W.T. (2021). “There’s Nothing Wrong With You”: Pain-Related Stigma in Adolescents With Chronic Pain. J. Pediatr. Psychol. 47, 456–468. 10.1093/jpepsy/jsab122.

29. Jackson, J.E. (2005). Stigma, liminality, and chronic pain: Mind-body borderlands. Am. Ethnol. 32, 332–353. 10.1525/ae.2005.32.3.332.

30. Wakefield, E.O., Kissi, A., Mulchan, S.S., Nelson, S., and Martin, S.R. (2022). Pain-related stigma as a social determinant of health in diverse pediatric pain populations. Front. Pain Res. 3.

31. Samulowitz, A., Gremyr, I., Eriksson, E., and Hensing, G. (2018). “Brave Men” and “Emotional Women”: A Theory-Guided Literature Review on Gender Bias in Health Care and Gendered Norms towards Patients with Chronic Pain. Pain Res. Manag. 2018, 6358624. 10.1155/2018/6358624.

32. Nijs, J., Lahousse, A., Kapreli, E., Bilika, P., Saraçoğlu, İ., Malfliet, A., Coppieters, I., De Baets, L., Leysen, L., Roose, E., et al. (2021). Nociplastic Pain Criteria or Recognition of Central Sensitization? Pain Phenotyping in the Past, Present and Future. J. Clin. Med. 10, 3203. 10.3390/jcm10153203.

33. Marcianò, G., Vocca, C., Evangelista, M., Palleria, C., Muraca, L., Galati, C., Monea, F., Sportiello, L., De Sarro, G., Capuano, A., et al. (2023). The Pharmacological Treatment of Chronic Pain: From Guidelines to Daily Clinical Practice. Pharmaceutics 15, 1165. 10.3390/pharmaceutics15041165.

34. Suri, P., Stanaway, I.B., Zhang, Y., Freidin, M.B., Tsepilov, Y.A., Carrell, D.S., Williams, F.M.K., Aulchenko, Y.S., Hakonarson, H., Namjou, B., et al. (2021). Genome-wide association studies of low back pain and lumbar spinal disorders using electronic health record data identify a locus associated with lumbar spinal stenosis. Pain Publish Ahead of Print. 10.1097/j.pain.0000000000002221.

35. Meng, W., Adams, M.J., Hebert, H.L., Deary, I.J., McIntosh, A.M., and Smith, B.H. (2018). A Genome-Wide Association Study Finds Genetic Associations with Broadly-Defined Headache in UK Biobank (N = 223,773). EBioMedicine 28, 180–186. 10.1016/j.ebiom.2018.01.023.

36. Jiang, L., Zheng, Z., Fang, H., and Yang, J. (2021). A generalized linear mixed model association tool for biobank-scale data. Nat. Genet. 53, 1616–1621. 10.1038/s41588-021-00954-4.

37. Eijsbouts, C., Zheng, T., Kennedy, N.A., Bonfiglio, F., Anderson, C.A., Moutsianas, L., Holliday, J., Shi, J., Shringarpure, S., 23andMe Research Team, et al. (2021). Genome-wide analysis of 53,400 people with irritable bowel syndrome highlights shared genetic pathways with mood and anxiety disorders. Nat. Genet. 53, 1543–1552. 10.1038/s41588-021-00950-8.

38. Hajdarevic, R., Lande, A., Mehlsen, J., Rydland, A., Sosa, D.D., Strand, E.B., Mella, O., Pociot, F., Fluge, Ø., Lie, B.A., et al. (2022). Genetic association study in myalgic encephalomyelitis/chronic fatigue syndrome (ME/CFS) identifies several potential risk loci. Brain. Behav. Immun. 102, 362–369. 10.1016/j.bbi.2022.03.010.

39. Grotzinger, A.D., Rhemtulla, M., de Vlaming, R., Ritchie, S.J., Mallard, T.T., Hill, W.D., Ip, H.F., Marioni, R.E., McIntosh, A.M., Deary, I.J., et al. (2019). Genomic structural equation modelling provides insights into the multivariate genetic architecture of complex traits. Nat. Hum. Behav. 3, 513–525. 10.1038/s41562-019-0566-x.

40. Bulik-Sullivan, B., Finucane, H.K., Anttila, V., Gusev, A., Day, F.R., Loh, P.-R., Duncan, L., Perry, J.R.B., Patterson, N., Robinson, E.B., et al. (2015). An atlas of genetic correlations across human diseases and traits. Nat. Genet. 47, 1236–1241. 10.1038/ng.3406.

41. 2.1 Calculating Sum of Effective Sample Size and Preparing GWAS Summary Statistics · GenomicSEM/GenomicSEM Wiki · GitHub https://github.com/GenomicSEM/GenomicSEM/wiki/2.1-Calculating-Sum-of-Effective-Sample-Size-and-Preparing-GWAS-Summary-Statistics.

42. Watanabe, K., Taskesen, E., van Bochoven, A., and Posthuma, D. (2017). Functional mapping and annotation of genetic associations with FUMA. Nat. Commun. 8, 1826. 10.1038/s41467-017-01261-5.

43. Bulik-Sullivan, B.K., Loh, P.-R., Finucane, H.K., Ripke, S., Yang, J., Patterson, N., Daly, M.J., Price, A.L., and Neale, B.M. (2015). LD Score regression distinguishes confounding from polygenicity in genome-wide association studies. Nat. Genet. 47, 291–295. 10.1038/ng.3211.

44. ten Klooster, P.M., de Graaf, N., and Vonkeman, H.E. (2019). Association between pain phenotype and disease activity in rheumatoid arthritis patients: a non-interventional, longitudinal cohort study. Arthritis Res. Ther. 21, 257. 10.1186/s13075-019-2042-4.

45. Cao, Y., Fan, D., and Yin, Y. (2020). Pain Mechanism in Rheumatoid Arthritis: From Cytokines to Central Sensitization. Mediators Inflamm. 2020, 2076328. 10.1155/2020/2076328.

46. Veluchamy, A., Hébert, H.L., van Zuydam, N.R., Pearson, E.R., Campbell, A., Hayward, C., Meng, W., McCarthy, M.I., Bennett, D.L.H., Palmer, C.N.A., et al. (2021). Association of Genetic Variant at Chromosome 12q23.1 With Neuropathic Pain Susceptibility. JAMA Netw. Open 4, e2136560. 10.1001/jamanetworkopen.2021.36560.

47. Ha, E., Bae, S.-C., and Kim, K. (2021). Large-scale meta-analysis across East Asian and European populations updated genetic architecture and variant-driven biology of rheumatoid arthritis, identifying 11 novel susceptibility loci. Ann. Rheum. Dis. 80, 558–565. 10.1136/annrheumdis-2020-219065.

48. Sollis, E., Mosaku, A., Abid, A., Buniello, A., Cerezo, M., Gil, L., Groza, T., Güneş, O., Hall, P., Hayhurst, J., et al. (2023). The NHGRI-EBI GWAS Catalog: knowledgebase and deposition resource. Nucleic Acids Res. 51, D977–D985. 10.1093/nar/gkac1010.

49. Gusev, A., Ko, A., Shi, H., Bhatia, G., Chung, W., Penninx, B.W.J.H., Jansen, R., de Geus, E.J., Boomsma, D.I., Wright, F.A., et al. (2016). Integrative approaches for large-scale transcriptome-wide association studies. Nat. Genet. 48, 245–252. 10.1038/ng.3506.

50. 50. The GTEx Consortium atlas of genetic regulatory effects across human tissues (2020). 15.

51. TWAS / FUSION http://gusevlab.org/projects/fusion/#download-pre-computed-predictive-models.

52. Kosek, E., Clauw, D., Nijs, J., Baron, R., Gilron, I., Harris, R.E., Mico, J.-A., Rice, A.S.C., and Sterling, M. (2021). Chronic nociplastic pain affecting the musculoskeletal system: clinical criteria and grading system. PAIN 162, 2629. 10.1097/j.pain.0000000000002324.

53. Bailly, F., Cantagrel, A., Bertin, P., Perrot, S., Thomas, T., Lansaman, T., Grange, L., Wendling, D., Dovico, C., and Trouvin, A.-P. (2020). Part of pain labelled neuropathic in rheumatic disease might be rather nociplastic. RMD Open 6, e001326. 10.1136/rmdopen-2020-001326.

54. Woods, R.H. (2022). Alopecia signals associated with calcitonin gene-related peptide inhibitors in the treatment or prophylaxis of migraine: A pharmacovigilance study. Pharmacother. J. Hum. Pharmacol. Drug Ther. 42, 758–767. 10.1002/phar.2725.

55. Aroke, E.N., and Powell-Roach, K.L. (2020). The Metabolomics of Chronic Pain Conditions: A Systematic Review. Biol. Res. Nurs. 22, 458–471. 10.1177/1099800420941105.

56. Boyd, J.T., LoCoco, P.M., Furr, A.R., Bendele, M.R., Tram, M., Li, Q., Chang, F.-M., Colley, M.E., Samenuk, G.M., Arris, D.A., et al. (2021). Elevated dietary ω-6 polyunsaturated fatty acids induce reversible peripheral nerve dysfunction that exacerbates comorbid pain conditions. Nat. Metab. 3, 762–773. 10.1038/s42255-021-00410-x.

57. Chiurchiù, V., Leuti, A., and Maccarrone, M. (2018). Bioactive Lipids and Chronic Inflammation: Managing the Fire Within. Front. Immunol. 9.

58. Li, S., Gao, D., and Jiang, Y. (2019). Function, Detection and Alteration of Acylcarnitine Metabolism in Hepatocellular Carcinoma. Metabolites 9, 36. 10.3390/metabo9020036.

59. Sanders, A.E., Weatherspoon, E.D., Ehrmann, B.M., Soma, P.S., Shaikh, S.R., Preisser, J.S., Ohrbach, R., Fillingim, R.B., and Slade, G.D. (2022). Circulating polyunsaturated fatty acids, pressure pain thresholds, and nociplastic pain conditions. Prostaglandins Leukot. Essent. Fatty Acids 184, 102476. 10.1016/j.plefa.2022.102476.

60. Navia-Pelaez, J.M., Choi, S.-H., dos Santos Aggum Capettini, L., Xia, Y., Gonen, A., Agatisa-Boyle, C., Delay, L., Gonçalves dos Santos, G., Catroli, G.F., Kim, J., et al. (2021). Normalization of cholesterol metabolism in spinal microglia alleviates neuropathic pain. J. Exp. Med. 218, e20202059. 10.1084/jem.20202059.

61. Robinson, G., Pineda-Torra, I., Ciurtin, C., and Jury, E.C. (2022). Lipid metabolism in autoimmune rheumatic disease: implications for modern and conventional therapies. J. Clin. Invest. 132. 10.1172/JCI148552.

62. Elma, Ö., Brain, K., and Dong, H.-J. (2022). The Importance of Nutrition as a Lifestyle Factor in Chronic Pain Management: A Narrative Review. J. Clin. Med. 11, 5950. 10.3390/jcm11195950.

63. Xiang, C., Chen, C., Li, X., Wu, Y., Xu, Q., Wen, L., Xiong, W., Liu, Y., Zhang, T., Dou, C., et al. (2022). Computational approach to decode the mechanism of curcuminoids against neuropathic pain. Comput. Biol. Med. 147, 105739. 10.1016/j.compbiomed.2022.105739.

64. Armstrong, C.W., McGregor, N.R., Butt, H.L., and Gooley, P.R. (2014). Chapter Five - Metabolism in Chronic Fatigue Syndrome. In Advances in Clinical Chemistry, G. S. Makowski, ed. (Elsevier), pp. 121–172. 10.1016/B978-0-12-801401-1.00005-0.

65. Hoel, F., Hoel, A., Pettersen, I.K.N., Rekeland, I.G., Risa, K., Alme, K., Sørland, K., Fosså, A., Lien, K., Herder, I., et al. A map of metabolic phenotypes in patients with myalgic encephalomyelitis/chronic fatigue syndrome. JCI Insight 6, e149217. 10.1172/jci.insight.149217.

66. Shelton, J.F., Shastri, A.J., Ye, C., Weldon, C.H., Filshtein-Sonmez, T., Coker, D., Symons, A., Esparza-Gordillo, J., Aslibekyan, S., and Auton, A. (2021). Trans-ancestry analysis reveals genetic and nongenetic associations with COVID-19 susceptibility and severity. Nat. Genet. 53, 801–808. 10.1038/s41588-021-00854-7.

67. Yang, Z., Zhou, D., Li, H., Cai, X., Liu, W., Wang, L., Chang, H., Li, M., and Xiao, X. (2020). The genome-wide risk alleles for psychiatric disorders at 3p21.1 show convergent effects on mRNA expression, cognitive function, and mushroom dendritic spine. Mol. Psychiatry 25, 48–66. 10.1038/s41380-019-0592-0.

68. Mei, Q., Li, X., Zhang, K., Wu, Z., Li, X., Meng, Y., Guo, M., Luo, G., Fu, X., and Han, W. (2017). Genetic and Methylation-Induced Loss of miR-181a2/181b2 within chr9q33.3 Facilitates Tumor Growth of Cervical Cancer through the PIK3R3/Akt/FoxO Signaling Pathway. Clin. Cancer Res. Off. J. Am. Assoc. Cancer Res. 23, 575–586. 10.1158/1078-0432.CCR-16-0303.

69. Ortiz, M., Towfic, F., Flynt, E., Stong, N., Lata, S., Sampath, D., Rozelle, D., Trotter, M., Corre, J., Avet-Loiseau, H., et al. (2019). Gene Expression and Genomic Markers Identify a Subpopulation of Poor Prognosis t(4;14) Patients in Newly Diagnosed Multiple Myeloma. Blood 134, 366. 10.1182/blood-2019-128907.

70. Gunther, K., Mowrey, K., and Farach, L.S. (2021). Two new reported cases of 16q22.3q23.3 duplication syndrome highlight intrafamilial variability and potential sex expression differences within a rare duplication syndrome. Clin. Case Rep. 9, 1629–1633. 10.1002/ccr3.3862.

71. Bozorgmehr, A., Sadeghi, B., Tabatabaei Zavari, E.S., Bahrami, E., Zamani, F., and Shahbazi, A. (2020). An integrative gene network-based approach to uncover the cellular and molecular infrastructures of schizophrenia. Life Sci. 260, 118345. 10.1016/j.lfs.2020.118345.

72. Ahmed, H., Akbari, H., Emami, A., and Akbari, M.R. (2017). Genetic Overview of Syndactyly and Polydactyly. Plast. Reconstr. Surg. Glob. Open 5, e1549. 10.1097/GOX.0000000000001549.

73. Li, Y.R., Glessner, J.T., Coe, B.P., Li, J., Mohebnasab, M., Chang, X., Connolly, J., Kao, C., Wei, Z., Bradfield, J., et al. (2020). Rare copy number variants in over 100,000 European ancestry subjects reveal multiple disease associations. Nat. Commun. 11, 255. 10.1038/s41467-019-13624-1.

74. Åkesson, K., Tenne, M., Gerdhem, P., Luthman, H., and McGuigan, F.E. (2015). Variation in the PTH2R gene is associated with age-related degenerative changes in the lumbar spine. J. Bone Miner. Metab. 33, 9–15. 10.1007/s00774-013-0550-x.

75. Littleton, S.H., Bradfield, J., Pippin, J.A., Su, C., Chesi, A., Wells, A.D., Berkowitz, R.I., Pahl, M.C., and Grant, S.F. (2022). 298-OR: Variant-to-Gene Mapping at the Childhood Obesity Locus on chr12q13 and Subsequent Luciferase Assay Analyses Implicate rs71329as a Causal Variant within the 3’ UTR of FAIM2. Diabetes 71, 298-OR. 10.2337/db22-298-OR.

76. Daya, M., Rafaels, N., Brunetti, T.M., Chavan, S., Levin, A.M., Shetty, A., Gignoux, C.R., Boorgula, M.P., Wojcik, G., Campbell, M., et al. (2019). Association study in African-admixed populations across the Americas recapitulates asthma risk loci in non-African populations. Nat. Commun. 10, 880. 10.1038/s41467-019-08469-7.

77. Demenais, F., Margaritte-Jeannin, P., Barnes, K.C., Cookson, W.O., Altmüller, J., Ang, W., Barr, R.G., Beaty, T.H., Becker, A.B., Beilby, J., et al. (2018). Multiancestry association study identifies new asthma risk loci that colocalize with immune cell enhancer marks. Nat. Genet. 50, 42–53. 10.1038/s41588-017-0014-7.

78. 78. CHARGE Inflammation working group, Ahluwalia, T.S., Prins, B.P., Abdollahi, M., Armstrong, N.J., Aslibekyan, S., Bain, L., Jefferis, B., Baumert, J., Beekman, M., et al. (2021). Genome-wide association study of circulating interleukin 6 levels identifies novel loci. Hum. Mol. Genet. 30, 393–409. 10.1093/hmg/ddab023.

79. Bergmans, R.S., Clauw, D.J., Flint, C., Harris, H., Lederman, S., and Schrepf, A. (2024). Chronic overlapping pain conditions increase the risk of long COVID features, regardless of acute COVID status. PAIN 165, 1112. 10.1097/j.pain.0000000000003110.

80. Wong, D., Auguste, G., Lino Cardenas, C.L., Turner, A.W., Chen, Y., Song, Y., Ma, L., Perry, R.N., Aherrahrou, R., Kuppusamy, M., et al. (2023). FHL5 Controls Vascular Disease-Associated Gene Programs in Smooth Muscle Cells. Circ. Res. 132, 1144–1161. 10.1161/CIRCRESAHA.122.321692.

81. O’Connor, E., Fourier, C., Ran, C., Sivakumar, P., Liesecke, F., Southgate, L., Harder, A.V.E., Vijfhuizen, L.S., Yip, J., Giffin, N., et al. (2021). Genome-Wide Association Study Identifies Risk Loci for Cluster Headache. Ann. Neurol. 90, 193–202. 10.1002/ana.26150.

82. 82. Atzeni, F., Cazzola, M., Benucci, M., Di Franco, M., Salaffi, F., and Sarzi-Puttini, P. (2011). Chronic widespread pain in the spectrum of rheumatological diseases. Best Pract. Res. Clin. Rheumatol. 25, 165–171. 10.1016/j.berh.2010.01.011.

83. Häuser, W., Sarzi-Puttini, P., and Fitzcharles, M.-A. (2019). Fibromyalgia syndrome: under-, over- and misdiagnosis. Clin. Exp. Rheumatol. 37, 90–97.

84. Häuser, W., Perrot, S., Sommer, C., Shir, Y., and Fitzcharles, M.-A. (2017). Diagnostic confounders of chronic widespread pain: not always fibromyalgia. Pain Rep. 2, e598. 10.1097/PR9.0000000000000598.

85. Richardson, A.C., Omar, M., Velarde, G., Missov, E., Percy, R., and Sattiraju, S. (2021). Right Atrial Appendage Thrombus in Atrial Fibrillation: A Case Report and Review of the Literature. J. Investig. Med. High Impact Case Rep. 9, 23247096211010048. 10.1177/23247096211010048.

86. 86. Regazzoli, D., Ancona, F., Trevisi, N., Guarracini, F., Radinovic, A., Oppizzi, M., Agricola, E., Marzi, A., Sora, N.C., Della Bella, P., et al. (2015). Left Atrial Appendage: Physiology, Pathology, and Role as a Therapeutic Target. BioMed Res. Int. 2015, 205013. 10.1155/2015/205013.

87. McIntyre, W.F., and Healey, J. (2017). Stroke Prevention for Patients with Atrial Fibrillation: Beyond the Guidelines. J. Atr. Fibrillation 9, 1475. 10.4022/jafib.1475.

88. 88. Peuckmann-Post, V., Eickhoff, R., Becker, M., and von der Laage, D. (2012). [ECG changes in patients with chronic non-cancer pain: a prospective observational study]. Schmerz Berl. Ger. 26, 419–424. 10.1007/s00482-012-1204-y.

89. Gong, C., Ding, Y., Liang, F., Wu, S., Tang, X., Ding, H., Huang, W., Yu, X., Zhou, L., Li, J., et al. (2022). Muscarinic receptor regulation of chronic pain-induced atrial fibrillation. Front. Cardiovasc. Med. 9, 934906. 10.3389/fcvm.2022.934906.

90. McIntyre, W.F., Connolly, S.J., and Healey, J.S. (2018). Atrial fibrillation occurring transiently with stress. Curr. Opin. Cardiol. 33, 58–65. 10.1097/HCO.0000000000000475.

91. McIntyre, W.F., Um, K.J., Cheung, C.C., Belley-Côté, E.P., Dingwall, O., Devereaux, P.J., Wong, J.A., Conen, D., Whitlock, R.P., Connolly, S.J., et al. (2019). Atrial fibrillation detected initially during acute medical illness: A systematic review. Eur. Heart J. Acute Cardiovasc. Care 8, 130–141. 10.1177/2048872618799748.

92. Rönnegård, A.-S., Nowak, C., Äng, B., and Ärnlöv, J. (2022). The association between short-term, chronic localized and chronic widespread pain and risk for cardiovascular disease in the UK Biobank. Eur. J. Prev. Cardiol. 29, 1994–2002. 10.1093/eurjpc/zwac127.

93. Di Carlo, M., Bianchi, B., Cipolletta, E., Farah, S., Filippucci, E., and Salaffi, F. Imaging of the peripheral nervous system in nociplastic pain: An ultrasound study in patients with fibromyalgia. J. Neuroimaging n/a. 10.1111/jon.13104.

94. Jo, Y.-S., Yoon, B., Hong, J.Y., Joung, C.-I., Kim, Y., and Na, S.-J. (2021). Tarsal tunnel syndrome in patients with fibromyalgia. Arch. Rheumatol. 36, 107–113. 10.46497/ArchRheumatol.2021.7952.

95. Xiang, H., Zhang, T., Al-Danakh, A., Yang, D., and Wang, L. (2022). Neuromodulation in Chronic Pelvic Pain: A Narrative Review. Pain Ther. 11, 789–816. 10.1007/s40122-022-00405-w.

