## Supplement for "Chronic Overlapping Pain Conditions and Nociplastic Pain"

**Supplemental Information**

**Table S1: Sources and sample sizes for COPC trait GWAS.**

| **Trait** | **Acronym** | **Source** | **Cases** | **Controls** | **Sample Prevalence** | **Population Prevalence** |
| --- | --- | --- | --- | --- | --- | --- |
| Chronic widespread pain | CWP | Rahman et al 2021 ^1^ | 6914 | 242929 | 0.029 | 0.01 ^2^ |
| Endometriosis | NA | FinnGenn (R9, May 2023) | 15088 | 107564 | 0.123 | 0.1 ^3^ |
| Low-back pain | LBP | Suri et al 2021 ^4^ | 49182 | 51629 | 0.488 | 0.075 ^5^ |
| Broad headache | NA | Meng et al 2018 ^6^ | 74461 | 149312 | 0.5 | 0.158 ^7^ |
| Temporomandibular joint disorder | TMJ | Jiang et al 2021 ^8^ (GWAS Catalog) | 217 | 456131 | 0.0005 | 0.3 ^9^ |
| Irritable Bowel Syndrome | IBS | Eijsbouts et al 2021 ^10^ (GWAS Catalog) | 53400 | 433201 | 0.12 | 0.1 ^11^ |

**Table S2:** Independent significant SNPs associated with pain traits in GWAS catalog searches with rsID as search terms.

| **GenomicLocus** | **rsID** | **p** | **Pain Trait Association** |
| --- | --- | --- | --- |
| 1 | rs1379837 | 4.78x10^-08^ | No |
| 1 | rs10804185 | 1.43x10^-08^ | No |
| 2 | rs653481 | 4.99x10^-09^ | No |
| 3 | rs529200 | 9.30x10^-10^ | No |
| 4 | rs138143060 | 1.33x10^-08^ | No |
| 5 | rs13208321 | 1.62x10^-11^ | No |
| 5 | rs10457146 | 2.98x10^-09^ | Migraine ^12^ |
| 5 | rs6941258 | 1.69x10^-10^ | Migraine ^12^ |
| 5 | rs2273621 | 2.64x10^-12^ | No |
| 6 | rs6961970 | 1.93x10^-08^ | No |
| 6 | rs10244257 | 9.86x10^-09^ | No |
| 7 | rs9918912 | 3.86x10^-10^ | No |
| 8 | rs10821159 | 1.46x10^-10^ | No |
| 9 | rs7905192 | 2.93x10^-09^ | No |
| 10 | rs7105462 | 8.27x10^-09^ | Back pain ^13^ |
| 11 | rs324014 | 2.54x10^-14^ | Itch (mosquito bite) ^14^ |
| 11 | rs11172113 | 3.94x10^-19^ | Migraine, headache ^6,15^ |
| 11 | rs7968719 | 4.85x10^-15^ | Migraine, headache ^12^ |
| 12 | rs2806954 | 4.15x10^-09^ | No |
| 12 | rs2813573 | 6.91x10^-09^ | No |
| 13 | rs2129316 | 1.15x10^-08^ | No |
| 13 | rs11850100 | 3.70x10^-08^ | No |
| 14 | rs6573501 | 2.65x10^-13^ | No |
| 15 | rs79724577 | 2.60x10^-08^ | No |

**Table S3:** Tissue-wide significant multivariate TWAS genes enriched in gene sets for GWAS catalog traits.

| GeneSet | P-value | adjusted P | genes |
| --- | --- | --- | --- |
| Cognitive function | 9.72x10-14 | 1.76x10^-10^ | RAB5B, SUOX, RPS26, ARHGAP27, PLEKHM1, LRRC37A4P, RHOA, TCTA, AMT, NICN1, CADM2 |
| Extremely high intelligence | 3.65x10^-12^ | 3.31x10^-09^ | GPX1, RHOA, TCTA, AMT, NICN1, DAG1, RNF123, GMPPB, IP6K1, CDHR4 |
| Sleep duration (short sleep) | 9.83x10^-12^ | 5.95x10^-09^ | GPX1, RHOA, TCTA, AMT, NICN1, DAG1, RNF123, GMPPB, IP6K1, FOXP2 |
| Headache | 2.51x10^-11^ | 1.10x10^-08^ | STAT6, LRP1, CRHR1, MAPT, PHACTR1, UFL1, FHL5 |
| Regular attendance at a religious group | 3.04x10^-11^ | 1.10x10^-08^ | RHOA, TCTA, AMT, NICN1, DAG1, RNF123, GMPPB, IP6K1, CDHR4 |
| Alcohol use disorder (total score) | 7.63x10^-09^ | 2.31x10^-06^ | PLEKHM1, DND1P1, CRHR1, MAPT-AS1, SPPL2C, MAPT |
| Mood instability | 1.01x10^-08^ | 2.59x10^-06^ | SORCS3, CRHR1, MAPT, GPX1, RHOA, NICN1, DAG1 |
| Neuroticism | 1.14x10^-08^ | 2.59x10^-06^ | MED19, ARHGAP27, PLEKHM1, CRHR1, SPPL2C, MAPT, EP300, L3MBTL2, FOXP2 |
| Migraine without aura | 2.30x10^-08^ | 4.63x10^-06^ | STAT6, LRP1, PHACTR1, UFL1, FHL5 |
| General factor of neuroticism | 1.91x10^-07^ | 3.47x10^-05^ | SORCS3, ARHGAP27, LRRC37A4P, CRHR1, MAPT-AS1, MAPT, CADM2 |
| General cognitive ability | 2.47x10^-07^ | 4.08x10^-05^ | AP1G1, ARHGAP27, MAPT-AS1, MAPT, RHOA, NICN1, IP6K1, CDHR4, CADM2 |
| Regular attendance at a gym or sports club | 2.70x10^-07^ | 4.08x10^-05^ | RNF123, GMPPB, IP6K1, CDHR4, CADM2 |
| Intelligence (MTAG) | 3.09x10^-07^ | 4.32x10^-05^ | SORCS3, SUOX, IST1, GPX1, NICN1, DAG1, IP6K1, CADM2, FAM120A, PHF2 |
| Multisite chronic pain | 4.45x10^-07^ | 5.77x10^-05^ | SORCS3, RNF123, GMPPB, C6orf106, FOXP2 |
| Medication use (anilides) | 2.00x10^-06^ | 2.42x10^-04^ | RABGAP1L, LRP1, MAPT |
| Asthma (adult onset) | 2.62x10^-06^ | 2.97x10^-04^ | RAB5B, SUOX, RPS26, NAB2, STAT6, LRP1 |
| Age at first birth | 8.65x10^-06^ | 9.24x10^-04^ | CRTC2, CREB3L4, RNF123, FOXP2 |
| Ability to confide in someone | 1.18x10^-05^ | 1.19x10^-03^ | FAM120AOS, FAM120A, PHF2 |
| Migraine - clinic-based | 1.62x10^-05^ | 1.55x10^-03^ | LRP1, PHACTR1, FHL5 |
| Migraine | 2.12x10^-05^ | 1.92x10^-03^ | STAT6, LRP1, PHACTR1, UFL1, FHL5 |
| Morning person | 2.82x10^-05^ | 2.33x10^-03^ | GLUL, TAT, MARVELD3, DHODH, RHOA, RNF123, PHACTR1 |
| Ulcerative colitis | 2.83x10^-05^ | 2.33x10^-03^ | GPX1, RHOA, TCTA, AMT, NICN1, DAG1, RNF123, GMPPB, IP6K1 |
| Idiopathic pulmonary fibrosis | 4.41x10^-05^ | 3.48x10^-03^ | CRHR1, SPPL2C, MAPT |
| Asthma or allergic disease (pleiotropy) | 4.65x10^-05^ | 3.52x10^-03^ | RAB5B, SUOX, RPS26, RDH16, NAB2, STAT6 |
| Hand grip strength | 4.84x10^-05^ | 3.52x10^-03^ | DHODH, SPPL2C, IP6K1, NLGN1, PHACTR1, UHRF1BP1 |
| Male-pattern baldness | 5.92x10^-05^ | 4.03x10^-03^ | FMNL1, ARHGAP27, PLEKHM1, CRHR1, SPPL2C, MAPT, LRRC37A2 |
| Crohn's disease | 6.00x10^-05^ | 4.03x10^-03^ | NRBP1, EP300, L3MBTL2, GPX1, RHOA, TCTA, AMT, NICN1, DAG1, RNF123, FOXP2 |
| Urate levels in obese individuals | 6.46x10^-05^ | 4.19x10^-03^ | LRFN5, CPS1, MPP6, FOXP2 |
| Parkinson's disease | 9.23x10^-05^ | 5.78x10^-03^ | ARHGAP27, PLEKHM1, CRHR1-IT1, CRHR1, SPPL2C, MAPT |
| Inflammatory bowel disease | 1.06x10^-04^ | 6.44x10^-03^ | NRBP1, PLAGL2, L3MBTL2, GPX1, RHOA, TCTA, AMT, NICN1, DAG1, RNF123, IP6K1 |
| Allergic disease (asthma, hay fever or eczema) | 1.63x10^-04^ | 9.57x10^-03^ | RAB5B, SUOX, RPS26, RDH16, NAB2, STAT6, ARHGAP27 |
| Intracranial volume | 3.25x10^-04^ | 1.84x10^-02^ | CRHR1, MAPT |
| Adventurousness | 3.49x10^-04^ | 1.92x10^-02^ | LRFN5, L3MBTL2, CADM2, NLGN1, FOXP2 |
| Drug experimentation measurement | 4.53x10^-04^ | 2.42x10^-02^ | CADM2, FOXP2 |
| Asthma (childhood onset) | 5.53x10^-04^ | 2.87x10^-02^ | RAB5B, SUOX, NAB2, STAT6, LRP1, ARHGAP27 |
| Amino acid levels | 6.03x10^-04^ | 3.04x10^-02^ | ACADL, CPS1 |
| Chronotype | 6.64x10^-04^ | 3.26x10^-02^ | SLC27A3, RABGAP1L, LRFN5, TAT, MARVELD3, CADM2, MED12L, PHACTR1, FOXP2 |
| Response to anti-TNF therapy in rheumatoid arthritis | 7.72x10^-04^ | 3.69x10^-02^ | ATXN1L, IST1 |
| General risk tolerance (MTAG) | 8.03x10^-04^ | 3.74x10^-02^ | SORCS3, LRFN5, EP300, L3MBTL2, CADM2, FOXP2 |
| Coronary artery disease | 8.53x10^-04^ | 3.87x10^-02^ | LRP1, RHOA, TCTA, AMT, PHACTR1, C6orf106, UHRF1BP1, FHL5 |
| Medication use (antimigraine preparations) | 9.62x10^-04^ | 4.12x10^-02^ | LRP1, PHACTR1 |
| Acylcarnitine levels | 9.62x10^-04^ | 4.12x10^-02^ | ACADL, CPS1 |
| Vitiligo | 9.77x10^-04^ | 4.12x10^-02^ | RAB5B, SUOX, RPS26, CBFA2T3 |
| Primary biliary cirrhosis | 1.02x10^-03^ | 4.19x10^-02^ | CRHR1, MAPT, PDGFB |

GeneSet = name of GWASCatalog gene set in FUMA.

**Table S4:** Tissue-wide significant multivariate TWAS genes enriched in additional gene sets.

| **GeneSet Category** | **GeneSet** | **P-value** | **adjusted P** | **genes** |
| --- | --- | --- | --- | --- |
| Chemical and Genetic Perturbation | SU_LIVER | 3.73x10^-06^ | 0.0123 | RDH16, TAT, DHODH, CPS1, SLC25A13 |
| KEGG | KEGG_NITROGEN_METABOLISM | 1.09x10^-04^ | 0.0202 | GLUL, CPS1, AMT |
| microRNA targets | AGCACTT_MIR93_MIR302A_MIR302B_MIR302C_MIR302D_MIR372_MIR373_MIR520E_MIR520A_MIR526B_MIR520B_MIR520C_MIR520D | 1.61x10^-05^ | 3.57x10^-03^ | CRTC2, PLEKHM1, PLAGL2, IP6K1, CADM2, MED12L, MFAP3L, PHF2, PPP6C |
| TF targets | GGGTGGRR_PAX4_03 | 6.32x10^-07^ | 3.85x10^-04^ | C1orf43, NAB2, STAT6, LRP1, LRFN5, AP1G1, CBFA2T3, FMNL1, MAPT, PDGFB, EP300, RHOA, TCTA, GMPPB, CADM2, ADD1, FAM120AOS, FAM120A, RABEPK |
| TF targets | EGR1_01 | 1.40x10^-06^ | 4.27x10^-04^ | HMGN2, CRTC2, NAB2, LRFN5, AP1G1, PDGFB, RHOA, TCTA, FOXP2 |
| TF targets | CTGCAGY_UNKNOWN | 6.06x10^-05^ | 0.0123 | HMGN2, RAB5B, LRFN5, CBFA2T3, MAPT, PDGFB, RHOA, TCTA, RNF123, CADM2, ATP2C1, SCAI |
| TF targets | PAX4_01 | 1.08x10^-04^ | 0.0164 | HMGN2, SORCS3, SUOX, LRP1, MAPT, PLAGL2, PDGFB |
| TF targets | AACTTT_UNKNOWN | 2.45x10^-04^ | 0.0255 | HMGN2, RABGAP1L, SORCS3, LRP1, HSPH1, CMTM4, CBFA2T3, FMNL1, PDGFB, GPX1, GMPPB, MED12L, P2RY13, NLGN1, C6orf106, UHRF1BP1, MPP6, FOXP2, SCAI |
| TF targets | TAANNYSGCG_UNKNOWN | 2.50x10^-04^ | 0.0255 | C1orf43, EP300, RHOA, TCTA |
| TF targets | RNGTGGGC_UNKNOWN | 3.27x10^-04^ | 0.0271 | HMGN2, NAB2, STAT6, LRP1, CBFA2T3, CRHR1, PDGFB, ST13, EP300, CADM2, ARPC5L |
| TF targets | TTCYRGAA_UNKNOWN | 4.52x10^-04^ | 0.0271 | HSPH1, MAPT, PDGFB, ST13, EP300, GPX1, ATP2C1 |
| TF targets | YGCGYRCGC_UNKNOWN | 4.81x10^-04^ | 0.0271 | C1orf43, SORCS3, LRFN5, MAPT, RHOA, TCTA, RNF123 |
| TF targets | YY1_01 | 4.89x10^-04^ | 0.0271 | AP1G1, IST1, RHOA, TCTA, SLC25A13, FOXP2 |
| TF targets | NGFIC_01 | 4.89x10^-04^ | 0.0271 | HMGN2, CRTC2, LRFN5, AP1G1, PDGFB, SCAI |
| TF targets | NFAT_Q4_01 | 7.01x10^-04^ | 0.0278 | PLAGL2, RHOA, TCTA, CADM2, FHL5, FOXP2 |
| TF targets | WTGAAAT_UNKNOWN | 7.62x10^-04^ | 0.0278 | C1orf43, AP1G1, CPS1, CADM2, P2RY13, P2RY12, SLC25A13, FOXP2, SCAI |
| TF targets | PPAR_DR1_Q2 | 7.68x10^-04^ | 0.0278 | LRP1, LRFN5, FMNL1, C6orf106, UHRF1BP1, SLC25A13 |
| TF targets | PAX2_02 | 7.85x10^-04^ | 0.0278 | MARVELD3, GPX1, RHOA, TCTA, MPP6, FOXP2 |
| TF targets | CEBPB_01 | 7.85x10^-04^ | 0.0278 | LRP1, AP1G1, EP300, GPX1, P2RY13, FOXP2 |
| TF targets | AP2_Q6 | 8.21x10^-04^ | 0.0278 | HMGN2, FMNL1, PLAGL2, PDGFB, EP300, PPP6C |
| TF targets | MMEF2_Q6 | 8.21x10^-04^ | 0.0278 | HMGN2, MYL6B, CPS1, RHOA, TCTA, FOXP2 |
| TF targets | EGR2_01 | 1.29x10^-03^ | 0.0415 | HMGN2, SORCS3, NAB2, PDGFB, SCAI |
| Computational | MODULE_294 | 1.05x10^-06^ | 8.98x10^-04^ | DHODH, ACADL, CPS1, AMT |
| Cancer gene modules | MODULE_294 | 1.05x10^-06^ | 4.51x10^-04^ | DHODH, ACADL, CPS1, AMT |

GeneSet = name of GWASCatalog gene set in FUMA. GeneSet Category = type of GeneSet.

**Table S5:** Specific tissue-wide significant multivariate TWAS genes enriched in additional gene sets.

| **GeneSet Category** | **GeneSet** | **P** | **P adj** | **genes** |
| --- | --- | --- | --- | --- |
| Chemical and genetic perturbation | SU_LIVER | 1.35x10^-06^ | 4.45x10^-03^ | RDH16, TAT, DHODH, CPS1, SLC25A13 |
| KEGG | KEGG_NITROGEN_METABOLISM | 5.89x10^-05^ | 0.0110 | GLUL, CPS1, AMT |
| microRNA targets | AGCACTT_MIR93_MIR302A_MIR302B_MIR302C_MIR302D_MIR372_MIR373_MIR520E_MIR520A_MIR526B_MIR520B_MIR520C_MIR520D | 2.53x10^-05^ | 5.59x10^-03^ | CRTC2, PLAGL2, IP6K1, CADM2, MED12L, MFAP3L, PHF2, PPP6C |
| TF targets | EGR1_01 | 2.40x10^-07^ | 1.46x10^-04^ | HMGN2, CRTC2, NAB2, LRFN5, AP1G1, PDGFB, RHOA, TCTA, FOXP2 |
| TF targets | GGGTGGRR_PAX4_03 | 1.40x10^-05^ | 4.27x10^-03^ | C1orf43, NAB2, LRFN5, AP1G1, CBFA2T3, PDGFB, EP300, RHOA, TCTA, GMPPB, CADM2, ADD1, FAM120AOS, FAM120A, RABEPK |
| TF targets | CTGCAGY_UNKNOWN | 4.06x10^-05^ | 8.26x10^-03^ | HMGN2, RAB5B, LRFN5, CBFA2T3, PDGFB, RHOA, TCTA, RNF123, CADM2, ATP2C1, SCAI |
| TF targets | TAANNYSGCG_UNKNOWN | 1.13x10^-04^ | 0.0125 | C1orf43, EP300, RHOA, TCTA |
| TF targets | AACTTT_UNKNOWN | 1.57x10^-04^ | 0.0125 | HMGN2, RABGAP1L, SORCS3, HSPH1, CMTM4, CBFA2T3, PDGFB, GPX1, GMPPB, MED12L, P2RY13, NLGN1, C6orf106, UHRF1BP1, MPP6, FOXP2, SCAI |
| TF targets | NGFIC_01 | 1.60x10^-04^ | 0.0125 | HMGN2, CRTC2, LRFN5, AP1G1, PDGFB, SCAI |
| TF targets | YY1_01 | 1.60x10^-04^ | 0.0125 | AP1G1, IST1, RHOA, TCTA, SLC25A13, FOXP2 |
| TF targets | WTGAAAT_UNKNOWN | 1.63x10^-04^ | 0.0125 | C1orf43, AP1G1, CPS1, CADM2, P2RY13, P2RY12, SLC25A13, FOXP2, SCAI |
| TF targets | PAX2_02 | 2.61x10^-04^ | 0.0167 | MARVELD3, GPX1, RHOA, TCTA, MPP6, FOXP2 |
| TF targets | MMEF2_Q6 | 2.73x10^-04^ | 0.0167 | HMGN2, MYL6B, CPS1, RHOA, TCTA, FOXP2 |
| TF targets | EGR2_01 | 5.09x10^-04^ | 0.0282 | HMGN2, SORCS3, NAB2, PDGFB, SCAI |
| TF targets | TCCCRNNRTGC_UNKNOWN | 8.00x10^-04^ | 0.0393 | MYL6B, HSPH1, PLAGL2, RHOA, TCTA |
| TF targets | TTCYRGAA_UNKNOWN | 8.82x10^-04^ | 0.0393 | HSPH1, PDGFB, ST13, EP300, GPX1, ATP2C1 |
| TF targets | YGCGYRCGC_UNKNOWN | 9.30x10^-04^ | 0.0393 | C1orf43, SORCS3, LRFN5, RHOA, TCTA, RNF123 |
| TF targets | GCANCTGNY_MYOD_Q6 | 9.84x10^-04^ | 0.0393 | HMGN2, RAB5B, LRFN5, CBFA2T3, PLAGL2, PDGFB, EP300, L3MBTL2, CADM2, FOXP2 |
| TF targets | CEBP_Q3 | 1.03x10^-03^ | 0.0393 | HMGN2, PDGFB, CADM2, MPP6, FOXP2 |
| TF targets | NFAT_Q6 | 1.10x10^-03^ | 0.0395 | PDGFB, GPX1, RHOA, TCTA, FOXP2 |
| TF targets | HSF2_01 | 1.18x10^-03^ | 0.0398 | HSPH1, CMTM4, CBFA2T3, PDGFB, ST13 |
| TF targets | AP4_Q6_01 | 1.60x10^-03^ | 0.0471 | HMGN2, RAB5B, CBFA2T3, PDGFB, CADM2 |
| TF targets | GATTGGY_NFY_Q6_01 | 1.65x10^-03^ | 0.0471 | HMGN2, RAB5B, IST1, LANCL1, L3MBTL2, RHOA, TCTA, GMPPB, CADM2, FAM120AOS, FAM120A |
| TF targets | NFAT_Q4_01 | 1.77x10^-03^ | 0.0471 | PLAGL2, RHOA, TCTA, CADM2, FOXP2 |
| TF targets | TAL1BETAITF2_01 | 1.80x10^-03^ | 0.0471 | MYL6B, CBFA2T3, PDGFB, CADM2, FOXP2 |
| TF targets | PAX4_01 | 1.91x10^-03^ | 0.0471 | HMGN2, SORCS3, SUOX, PLAGL2, PDGFB |
| TF targets | CEBPB_01 | 1.95x10^-03^ | 0.0471 | AP1G1, EP300, GPX1, P2RY13, FOXP2 |
| TF targets | AP2_Q6 | 2.02x10^-03^ | 0.0471 | HMGN2, PLAGL2, PDGFB, EP300, PPP6C |
| TF targets | NFY_Q6_01 | 2.02x10^-03^ | 0.0471 | HMGN2, RHOA, TCTA, CADM2, SLC25A13 |
| TF targets | EGR3_01 | 2.08x10^-03^ | 0.0471 | HMGN2, SORCS3, PDGFB |
| TF targets | TATAAA_TATA_01 | 2.27x10^-03^ | 0.0496 | HMGN2, RAB5B, LRFN5, PLAGL2, PDGFB, GPX1, RHOA, TCTA, P2RY12, FOXP2, RABEPK |
| Computational | MODULE_294 | 4.58x10^-07^ | 3.93x10^-04^ | DHODH, ACADL, CPS1, AMT |
| Cancer gene modules | MODULE_294 | 4.58x10^-07^ | 1.97x10^-04^ | DHODH, ACADL, CPS1, AMT |

GeneSet = name of GWASCatalog gene set in FUMA. GeneSet Category = type of GeneSet. Genes = overlapping genes

**Table S6:** Specific tissue-wide significant multivariate TWAS genes enriched in gene sets for GWAS catalog traits.

| **GeneSet** | **P-value** | **adjusted P** | **genes** |
| --- | --- | --- | --- |
| Extremely high intelligence | 4.42x10-13 | 8.02x10-10 | GPX1, RHOA, TCTA, AMT, NICN1, DAG1, RNF123, GMPPB, IP6K1, CDHR4 |
| Sleep duration (short sleep) | 1.20x10^-12^ | 1.09x10^-09^ | GPX1, RHOA, TCTA, AMT, NICN1, DAG1, RNF123, GMPPB, IP6K1, FOXP2 |
| Regular attendance at a religious group | 4.58x10^-12^ | 2.77x10^-09^ | RHOA, TCTA, AMT, NICN1, DAG1, RNF123, GMPPB, IP6K1, CDHR4 |
| Cognitive function | 6.79x10^-10^ | 3.08x10^-07^ | RAB5B, SUOX, RPS26, RHOA, TCTA, AMT, NICN1, CADM2 |
| Intelligence (MTAG) | 4.29x10^-08^ | 1.56x10^-05^ | SORCS3, SUOX, IST1, GPX1, NICN1, DAG1, IP6K1, CADM2, FAM120A, PHF2 |
| Regular attendance at a gym or sports club | 9.62x10^-08^ | 2.91x10^-05^ | RNF123, GMPPB, IP6K1, CDHR4, CADM2 |
| Multisite chronic pain | 1.59x10^-07^ | 4.12x10^-05^ | SORCS3, RNF123, GMPPB, C6orf106, FOXP2 |
| Mood instability | 2.84x10^-06^ | 6.45x10^-04^ | SORCS3, GPX1, RHOA, NICN1, DAG1 |
| Age at first birth | 3.82x10^-06^ | 7.70x10^-04^ | CRTC2, CREB3L4, RNF123, FOXP2 |
| Ulcerative colitis | 5.26x10^-06^ | 9.54x10^-04^ | GPX1, RHOA, TCTA, AMT, NICN1, DAG1, RNF123, GMPPB, IP6K1 |
| Ability to confide in someone | 6.38x10^-06^ | 1.05x10^-03^ | FAM120AOS, FAM120A, PHF2 |
| Crohn's disease | 8.43x10^-06^ | 1.28x10^-03^ | NRBP1, EP300, L3MBTL2, GPX1, RHOA, TCTA, AMT, NICN1, DAG1, RNF123, FOXP2 |
| Inflammatory bowel disease | 1.54x10^-05^ | 2.15x10^-03^ | NRBP1, PLAGL2, L3MBTL2, GPX1, RHOA, TCTA, AMT, NICN1, DAG1, RNF123, IP6K1 |
| Urate levels in obese individuals | 2.89x10^-05^ | 3.74x10^-03^ | LRFN5, CPS1, MPP6, FOXP2 |
| Morning person | 8.02x10^-05^ | 9.37x10^-03^ | GLUL, TAT, MARVELD3, DHODH, RHOA, RNF123 |
| General cognitive ability | 8.26x10^-05^ | 9.37x10^-03^ | AP1G1, RHOA, NICN1, IP6K1, CDHR4, CADM2 |
| Adventurousness | 1.33x10^-04^ | 0.0142 | LRFN5, L3MBTL2, CADM2, NLGN1, FOXP2 |
| Asthma or allergic disease (pleiotropy) | 1.83x10^-04^ | 0.0185 | RAB5B, SUOX, RPS26, RDH16, NAB2 |
| General risk tolerance (MTAG) | 2.67x10^-04^ | 0.0255 | SORCS3, LRFN5, EP300, L3MBTL2, CADM2, FOXP2 |
| Asthma (adult onset) | 2.92x10^-04^ | 0.0260 | RAB5B, SUOX, RPS26, NAB2 |
| Drug experimentation measurement | 3.01x10^-04^ | 0.0260 | CADM2, FOXP2 |
| Amino acid levels | 4.00x10^-04^ | 0.0330 | ACADL, CPS1 |
| Vitiligo | 4.51x10^-04^ | 0.0356 | RAB5B, SUOX, RPS26, CBFA2T3 |
| Cognitive ability, years of educational attainment or schizophrenia (pleiotropy) | 5.09x10^-04^ | 0.0373 | SORCS3, RPS26, CADM2, MPP6, FOXP2 |
| Response to anti-TNF therapy in rheumatoid arthritis | 5.13x10^-04^ | 0.0373 | ATXN1L, IST1 |
| Acylcarnitine levels | 6.40x10^-04^ | 0.0447 | ACADL, CPS1 |
| Anorexia nervosa | 7.31x10^-04^ | 0.0476 | RAB5B, SUOX, RPS26 |
| Chronotype | 7.35x10^-04^ | 0.0476 | SLC27A3, RABGAP1L, LRFN5, TAT, MARVELD3, CADM2, MED12L, FOXP2 |

GeneSet = name of GWASCatalog gene set in FUMA. Genes = overlapping genes.

**Table S7:** Non-specific tissue-wide significant multivariate TWAS genes enriched in additional gene sets.

| **GeneSet Category** | **GeneSet** | **P-value** | **adjusted P** | **genes** |
| --- | --- | --- | --- | --- |
| Cancer gene modules | GNF2_PAK2 | 1.27x10^-04^ | 0.0325 | STAT6, FMNL1 |
| Cancer gene modules | GNF2_ICAM3 | 2.10x10^-04^ | 0.0325 | STAT6, FMNL1 |
| Cancer gene modules | GNF2_PTPN6 | 2.66x10^-04^ | 0.0325 | STAT6, FMNL1 |
| Cancer gene modules | GNF2_CD53 | 3.46x10^-04^ | 0.0325 | STAT6, FMNL1 |
| Cancer gene modules | GNF2_SELL | 3.81x10^-04^ | 0.0325 | STAT6, FMNL1 |
| Cancer gene modules | GNF2_MYD88 | 6.01x10^-04^ | 0.0428 | STAT6, FMNL1 |

GeneSet = name of GWASCatalog gene set in FUMA. GeneSet Category = type of GeneSet. Genes = overlapping genes.

**Table S8:** Non-specific tissue-wide significant multivariate TWAS genes enriched in gene sets for GWAS catalog traits.

| **GeneSet** | **P-value** | **adjusted P** | **genes** |
| --- | --- | --- | --- |
| Migraine without aura | 5.13x10^-10^ | 9.31x10^-07^ | STAT6, PHACTR1, UFL1, FHL5 |
| Headache | 1.97x10^-09^ | 1.79x10^-06^ | STAT6, PHACTR1, UFL1, FHL5 |
| Migraine | 1.26x10^-07^ | 7.63x10^-05^ | STAT6, PHACTR1, UFL1, FHL5 |
| Alcohol use disorder (total score) | 1.22x10^-06^ | 5.55x10^-04^ | DND1P1, MAPT-AS1, SPPL2C |
| Male-pattern baldness | 9.50x10^-06^ | 3.45x10^-03^ | FMNL1, ARHGAP27, SPPL2C, LRRC37A2 |
| Migraine - clinic-based | 2.35x10^-05^ | 6.86x10^-03^ | PHACTR1, FHL5 |
| General factor of neuroticism | 2.65x10^-05^ | 6.86x10^-03^ | ARHGAP27, LRRC37A4P, MAPT-AS1 |
| Sense of smell | 6.51x10^-05^ | 0.0148 | LRRC37A4P, LRRC37A2 |
| Parkinson's disease | 1.43x10^-04^ | 0.0288 | ARHGAP27, CRHR1-IT1, SPPL2C |

GeneSet = name of GWASCatalog gene set in FUMA. GeneSet Category = type of GeneSet. Genes = overlapping genes.

**Table S9:** Fisher’s exact tests of counts for ICD10 codes comparing MCP controls and cases in UK Biobank

| **Trait** | **N MCP case with Trait** | **N MCP case no Trait** | **N MCP control with Trait** | **N MCP control without Trait** | **Odds** | **CI (L)** | **CI (U)** | **P** |
| --- | --- | --- | --- | --- | --- | --- | --- | --- |
| Dorsalgia (Back pain) | 34425 | 161709 | 25066 | 217467 | 1.85 | 1.81 | 1.88 | << 9.76x10-302 |
| Post-viral fatigue | 2665 | 193469 | 1906 | 240627 | 1.74 | 1.64 | 1.85 | 1.85x10-76 |
| Migraine | 17952 | 178182 | 11308 | 231225 | 2.06 | 2.01 | 2.11 | << 9.76x10-302 |
| IBS | 13955 | 182179 | 9277 | 233256 | 1.93 | 1.87 | 1.98 | << 9.76x10-302 |
| Endometriosis | 1430 | 194704 | 999 | 241534 | 1.78 | 1.64 | 1.93 | 1.07x10-44 |
| Tension Headache | 2576 | 193558 | 1566 | 240967 | 2.05 | 1.92 | 2.18 | 6.42x10^-11^4 |
| Interstitial Cystitis | 306 | 195828 | 173 | 242360 | 2.19 | 1.81 | 2.65 | 3.74x10-17 |
| Fibromyalgia | 2288 | 193846 | 638 | 241895 | 4.48 | 4.10 | 4.89 | 9.76x10-302 |
| Rheumatoid arthritis | 5509 | 190625 | 3015 | 239518 | 2.29 | 2.19 | 2.4 | 8.5x10-305 |

Case = any non-zero multisite chronic pain (MCP) trait value, Control = MCP trait value 0. CI (L) = confidence interval of odds ratio (lower bound value), CI (U) = confidence interval (upper bound value).

Table S10: Liability scale SNP-heritability of each chronic overlapping pain condition (COPC) trait.

| **Trait** | **SNP-h2** | **SE** |
| --- | --- | --- |
| CWP | 0.167 | 0.0142 |
| IBS | 0.077 | 0.0045 |
| Headache | 0.107 | 0.0053 |
| LBP | 0.0138 | 0.0021 |
| TMJ | 0.1726 | 0.73 |
| Endometriosis | 0.0508 | 0.01 |

SNP-h2 = SNP-heritability value (calculated using LD-score regression). SE = standard error of SNP-h2 value.

**References**

1.	Rahman, M.S., Winsvold, B.S., Chavez Chavez, S.O., Børte, S., Tsepilov, Y.A., Sharapov, S.Z., HUNT All-In Pain, Aulchenko, Y.S., Hagen, K., Fors, E.A., et al. (2021). Genome-wide association study identifies *RNF123* locus as associated with chronic widespread musculoskeletal pain. Ann. Rheum. Dis. *80*, 1227–1235. 10.1136/annrheumdis-2020-219624.

2.	Andrews, P., Steultjens, M., and Riskowski, J. (2018). Chronic widespread pain prevalence in the general population: A systematic review. Eur. J. Pain *22*, 5–18. 10.1002/ejp.1090.

3.	Taylor, H.S., Kotlyar, A.M., and Flores, V.A. (2021). Endometriosis is a chronic systemic disease: clinical challenges and novel innovations. The Lancet *397*, 839–852. 10.1016/S0140-6736(21)00389-5.

4.	Suri, P., Stanaway, I.B., Zhang, Y., Freidin, M.B., Tsepilov, Y.A., Carrell, D.S., Williams, F.M.K., Aulchenko, Y.S., Hakonarson, H., Namjou, B., et al. (2021). Genome-wide association studies of low back pain and lumbar spinal disorders using electronic health record data identify a locus associated with lumbar spinal stenosis. Pain *Publish Ahead of Print*. 10.1097/j.pain.0000000000002221.

5.	Wu, A., March, L., Zheng, X., Huang, J., Wang, X., Zhao, J., Blyth, F.M., Smith, E., Buchbinder, R., and Hoy, D. (2020). Global low back pain prevalence and years lived with disability from 1990 to 2017: estimates from the Global Burden of Disease Study 2017. Ann. Transl. Med. *8*, 299–299. 10.21037/atm.2020.02.175.

6.	Meng, W., Adams, M.J., Hebert, H.L., Deary, I.J., McIntosh, A.M., and Smith, B.H. (2018). A Genome-Wide Association Study Finds Genetic Associations with Broadly-Defined Headache in UK Biobank (N = 223,773). EBioMedicine *28*, 180–186. 10.1016/j.ebiom.2018.01.023.

7.	Stovner, L.J., Hagen, K., Linde, M., and Steiner, T.J. (2022). The global prevalence of headache: an update, with analysis of the influences of methodological factors on prevalence estimates. J. Headache Pain *23*, 34. 10.1186/s10194-022-01402-2.

8.	Jiang, L., Zheng, Z., Fang, H., and Yang, J. (2021). A generalized linear mixed model association tool for biobank-scale data. Nat. Genet. *53*, 1616–1621. 10.1038/s41588-021-00954-4.

9.	Valesan, L.F., Da-Cas, C.D., Réus, J.C., Denardin, A.C.S., Garanhani, R.R., Bonotto, D., Januzzi, E., and de Souza, B.D.M. (2021). Prevalence of temporomandibular joint disorders: a systematic review and meta-analysis. Clin. Oral Investig. *25*, 441–453. 10.1007/s00784-020-03710-w.

10.	Eijsbouts, C., Zheng, T., Kennedy, N.A., Bonfiglio, F., Anderson, C.A., Moutsianas, L., Holliday, J., Shi, J., Shringarpure, S., 23andMe Research Team, et al. (2021). Genome-wide analysis of 53,400 people with irritable bowel syndrome highlights shared genetic pathways with mood and anxiety disorders. Nat. Genet. *53*, 1543–1552. 10.1038/s41588-021-00950-8.

11.	Canavan, C., West, J., and Card, T. (2014). The epidemiology of irritable bowel syndrome. Clin. Epidemiol. *6*, 71–80. 10.2147/CLEP.S40245.

12.	Meng, W., Reel, P.S., Nangia, C., Rajendrakumar, A.L., Hebert, H.L., Guo, Q., Adams, M.J., Zheng, H., Lu, Z.H., 23andMe Research Team, et al. (2023). A Meta-Analysis of the Genome-Wide Association Studies on Two Genetically Correlated Phenotypes Suggests Four New Risk Loci for Headaches. Phenomics Cham Switz. *3*, 64–76. 10.1007/s43657-022-00078-7.

13.	Freidin, M.B., Tsepilov, Y.A., Palmer, M., Karssen, L.C., Suri, P., Aulchenko, Y.S., Williams, F.M.K., and CHARGE Musculoskeletal Working Group (2019). Insight into the genetic architecture of back pain and its risk factors from a study of 509,000 individuals. Pain *160*, 1361–1373. 10.1097/j.pain.0000000000001514.

14.	Jones, A.V., Tilley, M., Gutteridge, A., Hyde, C., Nagle, M., Ziemek, D., Gorman, D., Fauman, E.B., Chen, X., Miller, M.R., et al. (2017). GWAS of self-reported mosquito bite size, itch intensity and attractiveness to mosquitoes implicates immune-related predisposition loci. Hum. Mol. Genet. *26*, 1391–1406. 10.1093/hmg/ddx036.

15.	Chasman, D.I., Schürks, M., Anttila, V., de Vries, B., Schminke, U., Launer, L.J., Terwindt, G.M., van den Maagdenberg, A.M.J.M., Fendrich, K., Völzke, H., et al. (2011). Genome-wide association study reveals three susceptibility loci for common migraine in the general population. Nat. Genet. *43*, 695–698. 10.1038/ng.856.
